# A reproducible MRI-to-FE workflow for generating population-specific and subject-specific finite element head models

**DOI:** 10.64898/2026.04.28.26351900

**Authors:** Christian John A. Saludar, Maryam Tayebi, Eryn Kwon, Jacob Mathew, Joshua McGeown, William Schierding, Mātai mTBI Research Group, Alan Wang, Justin Fernandez, Samantha Holdsworth, Vickie Shim

**Author notes:** Corresponding Author: Vickie Shim, Auckland Bioengineering Institute, University of Auckland, 70 Symonds Street, Auckland, New Zealand.

## Abstract

**Purpose:** Finite element (FE) models are widely used to study traumatic brain injury, but most are based on single-subject scans or generic atlases that may not represent specific cohorts, introducing variability in biomechanical outputs. This study presents a reproducible framework for generating a cohort-specific MRI-derived template brain and constructing a representative FE head model.

**Methods:** A cohort-specific template was created from T1-weighted and diffusion-weighted MRI scans of collision sport athletes (n = 78). An FE head model was then generated from this template, with material properties mapped using MRI-derived microstructural information. The anatomical accuracy of the template was evaluated using deformation-based morphometry against standard brain atlases. The FE model was validated against cadaveric head impact experiments. A subject-specific FE model was also developed to test the pipeline’s applicability and model performance.

**Results:** Registration of an external non-collision sport controls (n = 20) to the template showed reduced deformation compared with registration to the MNI152 and an existing adolescent atlas, indicating improved cohort representation. Validation against cadaveric data demonstrated small to moderate correlations between simulated and experimental nodal displacements. Subject-specific and fiber dispersion-informed models showed significantly different strain distributions compared with isotropic models under low-intensity impacts, but no significant differences under high-intensity impacts.

**Conclusion:** The proposed framework enables reproducible construction of cohort-specific FE brain models using open-access tools. Results highlight the importance of subject-and cohort-specific modelling for accurately characterizing brain biomechanics. This approach improves the anatomical and microstructural realism of FE models and supports more precise mapping of mechanical strain to clinical outcomes in TBI.

**STATEMENTS AND DECLARATIONS:** We declare that we have no financial or personal relationships with individuals, companies, or organisations that could inappropriately influence the content of this work. We also confirm that we hold no professional or personal interests in any product, service, or company that could be perceived as affecting the content and quality of this manuscript.

## INTRODUCTION

Over the past decades, research has experienced exponential growth in the use of finite element (FE) head models to understand brain biomechanics of traumatic brain injury (TBI). Advances in computational methods and neuroimaging have enabled increasingly sophisticated models that incorporate detailed representations of complex brain structures [1,2]. In particular, the use of magnetic resonance imaging (MRI) has been significant in providing anatomical structures to guide FE model development. However, these models are mostly derived from a single-subject scan [3], the 50^th^ percentile of a cohort[4], or a general open-source brain atlas [5]. In addition, while MRI has provided anatomical details, these were based on subjects from a particular age or sex group, leading to geometric differences when applied to other cohorts. This highlights the need to better represent the anatomical characteristics of the populations to which they are applied.

This is particularly important because brain geometry varies substantially across age, sex, and developmental stage, and these differences have been shown to influence the brain’s biomechanical response during head impacts [6–8]. Consequently, FE models derived from non-representative templates may introduce geometric bias, which may reduce the validity of simulation results. Moreover, current adult human brain atlases may not be best suited for studying specific populations, particularly given the rapidly developing nature of the brain in paediatrics and adolescents [9]. To address this, population-specific MRI atlases have been proposed as anatomically representative templates for targeted cohorts. For example, Zou et al. (2021) developed an MRI-derived adolescent brain atlas for collision-sport athletes to reduce registration bias by using a population-specific atlas template, as mismatch between the template and study population can confound neuroimaging findings [9].

In addition to the importance of a population-specific atlas, studies have highlighted the need to develop subject-specific FE head models [7]. FE models have been limited in the ability to efficiently generate subject-specific models that accurately capture an individual’s unique brain geometry and material properties[10]. While sophisticated models have emerged and studies have developed multiple subject models, they mostly used homogeneous brain materials, which do not account for the heterogeneity and subject-specificity of brain material properties. In our previous work, Shim et al. (2022) developed a computational pipeline in an ovine model that generates a subject-specific FE model from advanced MRI scans. Although this was one of the first fully subject-specific FE models of the sheep brain predicting structural alteration after an induced impact, its application and usability for human brain analysis were identified as future directions of the paper.

The necessity of these anatomically accurate FE head models is most evident in the study of TBI, a global public health problem affecting 70 million individuals annually worldwide [11]. It causes significant socio-economic problems[12], yet its mechanism remains poorly understood. Mild TBI (mTBI) accounts for up to 95% of all TBI cases[13]. These are caused by external biomechanically plausible mechanisms of injury involving direct or indirect forces transmitted to the head[14] and are followed by clinical signs of physiological disruption of brain function, worsened symptoms within 72 hours post-injury, and impairment in cognitive, balance, or oculomotor function. In addition to the high prevalence of TBI and mTBI, interest has grown in exposure to sports-related head acceleration events (HAE). While many HAEs are subclinical and therefore do not produce overt acute clinical symptoms (*i.e.,* not TBI or mTBI), they have been shown to induce long-term neurological effects [15]. However, some high-intensity HAEs cross the clinical threshold causing overt acute clinical symptoms (*i.e.,* an mTBI incident). The short-and long-term effects of mTBI and subclinical HAE on brain health remain unclear, prompting growing interest[16–19].

Neuroimaging studies have examined neurological and microstructural alterations associated with TBI, mTBI, and repetitive HAE, as well as their relationships with head-impact kinematics measured by instrumented tools. However, findings remain heterogenous, potentially driven by simplified quantification of impact exposure that rely mainly on linear and rotational acceleration[20,21]. Properly quantifying how energy dissipates from these mTBI or HAE events into neuronal tissue and structures is critical for understanding their underlying mechanisms [21]. By modelling these dynamics, researchers can better characterise the mechanical behaviour of specific brain regions and map those most strained during an impact [20]. This biomechanical mapping influences the resulting pattern of structural damage and can suggest specific neurological sequelae[22]. Ultimately, advancing toward such precise quantification could support improved diagnosis and prognosis, with the potential to tailor therapy and rehabilitation to each subject’s specific injury profile.

Building on prior studies, the purpose of this work is to develop a population-specific brain template and atlas for adolescent collision-sport athletes, create an FE head model from this template, and validate the model against existing human cadaveric data. The overarching goal is to establish a reproducible MRI-to-FE pipeline that enables research groups to personalise their FE models for their specific cohort of interest. This pipeline integrates open-source packages across the full workflow, from MRI pre-processing to FE simulation, enabling reproducibility across research groups. Furthermore, the existing subject-specific FE model generation pipeline will be applied to the human brain template developed, allowing adaptation of the population template to subject scans and assignment of subject-specific material properties. Finally, a comparison of brain responses between the template-based and a subject-specific FE head model is performed as a proof-of-concept for generating subject-specific models using the proposed pipeline.

## METHODS

### Ethics approval

This work was conducted under ethical approval from the New Zealand Health and Disability Ethics Committee (HDEC 20/NTB/14) in accordance with the Declaration of Helsinki. Participants aged >16 years old provided written informed consent, while those <16 years old provided written assent with parental consent.

### Participants and design

#### Subject recruitment

This work is part of an ongoing longitudinal study in New Zealand recruiting adolescent male rugby union athletes across multiple competitive secondary school seasons[16,23]. Potential participants were excluded if they had a pre-existing diagnosed neuropsychiatric condition, had dental braces that could cause imaging artefacts, or had a self-reported mTBI within six months prior to the study season of participation. mTBI diagnosis was defined based on the WHO Neurotrauma Task Force [24] and the American Congress of Rehabilitation Medicine [14]. Participants meeting inclusion criteria completed a multimodal MRI scanning at early-, mid-, and post-season. The term early-season is used rather than pre-season because many players participate in a full collision training camp to determine which team they will be part of (1st or 2nd XV) prior to the baseline (early-season) scanning. For this work, 78 rugby players compose our collision sport group from whom the template was created.

#### Mouthguard instrumentation

Each participant was provided with an instrumented mouthguard, custom-moulded from their dental scan, to record HAE exposure throughout the season. HitIQ® Nexus A9 was used in 2021, and Prevent Biometrics Instant Custom Fit 2.0 in 2023. The mouthguard had been previously validated against reference sensors in an aluminium headform across a range of impact intensities[25], as well as against on-field video validation[26]. Recorded kinematic data, triggered and processed through each company’s signal processing pipeline, were exported from the respective iMG portals. No additional filtering or verification was performed. HAE recorded using these mouthguards will serve as input for comparing non-mTBI and mTBI-causing HAE.

#### MRI acquisition

All MR scans were acquired using a 3.0 T MR Scanner (GE SIGNA Premier; General Electric, MI, USA) equipped with a 48-channel head coil. A high-resolution 3D T1-weighted inversion recovery (IR)-prep, fast IR RF-Spoiled Gradient echo sequence (repetition time [TR] = 6.6 ms, echo time [TE] = 2.7 ms, flip angle = 12°, inversion time [TI] = 600 ms, matrix size = 512 × 512, FOV = 224 × 224 mm, number of slices = 320, voxel size = 0.4 × 0.4 × 0.5 mm) was acquired. A multi-shell spin-echo diffusion-weighted MRI was also performed (TR = 4500 ms, TE = 70 ms, flip angle = 90°, matrix size = 128 × 128, FOV = 260 × 260 mm, number of slices = 80, voxel size = 2 × 2 × 2 mm, parallel reduction factor = 2, multiband factor = 3, b-value = 0, 1000, 2000, 3000 s/mm2, 54 gradient directions = 4, 15, 15, 20, respectively). The current analysis is limited to T1-W and diffusion sequences with a total scan time of 10 min.

### MRI Pre-processing

#### Pre-processing

For the template generation, only the early-season scans were included. This is to control for any microstructural alteration due to repetitive subclinical HAE exposure throughout the season. All diffusion MRI (dMRI) and T1-weighted structural images were manually reviewed in FSLeyes to identify significant head motion, imaging artefacts, or missing slices. Severely affected scans were excluded prior to pre-processing.

#### Structural image

T1-weighted images were skull-stripped using HD-Bet[27]. Following extraction, the brain images were segmented into three tissue classes – Gray Matter (GM), White Matter (WM), and Cerebrospinal Fluid (CSF) – using CAT[28]. These were used as a priori for N4 bias field inhomogeneity correction using ANTs[29].

#### Diffusion MRI

dMRI were pre-processed using FSL [30,31] and MRtrix3[32] to remove noise [33], correct B1-field inhomogeneity, remove Gibbs’ ringing artefacts, and correct eddy current and motion artefacts. Eddy-current distortion was corrected using FSL’s ‘eddy’ function [34] with slice-to-volume correction. The Synb0-DISCO [35] was used for EPI distortion correction and field map estimation. After pre-processing, quality control was performed using Eddy QUAD [36] and by visual inspection in FSLeyes. Images with persistent artefacts were excluded from further processing. Diffusion tensor imaging metrics were derived using FSL(*dtifit*)[37]. In this pipeline, only the fractional anisotropy (FA) map is included.

### Study-specific MRI template and atlas generation

The image-processing pipeline was designed to generate a study-specific anatomical reference and to transfer MRI-derived tissue and fibre information into a finite element model. The term “template” refers to the study-specific common reference space generated from the multimodal MRI data, primarily the co-registered T1-weighted and FA images. Within this common space, a separate population fibre orientation distributions (FOD) template was generated to represent average white matter fibre orientation information for FE modelling. The term atlas refers to the labelled version of this template after anatomical parcellations, tissue labels, and white matter tract masks were incorporated. In this study, the labelled template was referred to as the “ABI-Matai Adolescent Rugby Atlas 78” or “AMARA-78” Prior to template construction, each subject’s FA image was co-registered to the corresponding T1-weighted image via rigid registration (FSL *flirt*). A multimodal population template was then generated using ANTS *antsMultivariateTemplateConstruction.sh*, with both T1-weighted and FA images used to guide registration [38]. The template was initialised using the 1mm isotropic MNI152 space as a common reference. Each subject image was first rigidly aligned to MNI152 and averaged to create an initial template. Each subject’s image was non-linearly registered to the initial template using Greedy SyN *(-t GR*) transformation model including cross-correlation for within-modality alignment *(-s CC*) and mutual information where intermodal alignment was required. The resulting warped images were then averaged across participants to create an updated template. This process was repeated iteratively, with each new template used as the target for the next registration step. Iteration continued until the Pearson cross-correlation similarity index between subsequent template iterations reached a plateau for both the T1-weighted and FA templates.

### Population FOD template generation

Each participant’s FA image was affine registered to the template FA (*antsRegistration.sh*), and the resulting transformation was applied to the pre-processed diffusion-weighted images (*antsApplyTransforms*). The b-vectors were rotated to account for the spatial transformation. Given that white matter fibre tract orientation can significantly influence tissue deformation along the principal fibre direction [39], the fibre orientation distributions (FOD) were estimated for each participant.

Response functions were estimated using the ‘*dhollander’* algorithm [40], followed by multi-shell multi-tissue constrained spherical deconvolution (MSMT-CSD) in MRTrix3. Output FODs were then used as input to the ‘population_template’ in MRTrix3 to generate a study-specific FOD template. From this template, the first three FOD peaks were extracted and used to derive the fibre direction vectors for subsequent FE modelling.

### Atlas and ROI generation

The template T1-weighted structural image was parcellated and segmented using ‘SynthSeg’[41]. This outputs a label map with regions indexed as outlined in Supplementary Material 1 following the Desikan-Killiany parcellation[42]. To parcellate the WM regions, the template FOD peaks served as input to TractSeg [43]. The GM parcellation, tissue segmentations, and WM tract masks were incorporated into the population template to create the labelled AMARA-78 atlas. This atlas provided the anatomical reference used to localise regional brain responses and guide MRI-to-FE material assignments.

### AMARA-78 MRI evaluation

AMARA-78 T1 template was evaluated using a cohort of 20 male non-collision-sport athletes assessed at a single time point, who served as controls in our longitudinal study set up [16,23]. Participants were actively participated in non-collision sports including basketball, track and field, rowing, and waka ama (outrigger canoe racing). MRI scans were acquired using the same protocol as for the collision-sport athletes.

To assess potential registration bias, deformation-based morphometry was performed using an approach similar to Zou et al.[9]. Pre-processed T1-weighted images were normalised separately to the MNI152-1mm template, Purdue Neurotrauma Group (PNG) [9] and the AMARA-78-T1 template. The MNI152-1mm template is derived from 152 young adult brains and averaged together after high-dimensional nonlinear registration into the common MNI152 coordinate system. On the other hand, PNG is a brain atlas derived from early-to-middle adolescent collision-sport athletes.

For each registration, the logarithm of Jacobian determinant (logJ), representing local volume difference, was estimated in ANTs. The absolute logJ (|logJ|) values were computed and transformed to the MNI152 space to ensure all deformation maps were aligned on a common voxel grid for voxel-wise statistical comparison. Voxel-wise paired-sample t-tests were then performed in FSL randomise with 5000 permutations, using threshold-free cluster enhancement and family-wise error correction [44]. Voxels were considered statistically significant at a threshold of *p* < 0.05.

### FE model generation

A surface mesh was generated from the AMARA-78 T1 derived brain mask using 3DSlicer[45]. To represent anatomical continuity with the spinal cord, the lower end of the brainstem was subsequently extruded to include a simplified spinal cord segment. The resulting STL file was then imported into PolyCube (https://www.cs.ubc.ca/labs/imager/tr/2018/HexDemo/), which automatically generated a hexahedral volume mesh that is compatible with direct import into FEBio as a template FE model [46]. Afterwards an in-house Python script established element-voxel correspondence by identifying the MRI voxels contained within that element. The voxel values were then aggregated to assign element-wise properties, enabling MRI-guided assignment of tissue class and material parameters to the FE mesh. Thus, we first mapped MRI-derived segmentations onto the FE mesh, assigning each element to tissue classes including GM, WM, and CSF. As for WM, spatially varying heterogeneous material properties were assigned by linking each element to corresponding DTI-derived voxel information. This approach has previously been used to assign element-specific material information in different FE modelling applications [10,21,47]. The falx cerebri and tentorium cerebelli, which are important intracranial structures [5], were represented as manually defined single-layer element structures positioned between the cerebral hemispheres and between the cerebrum and cerebellum, respectively.

Brain tissue was modelled using a hyper-viscoelastic fibre-reinforced anisotropic model based on Gasser, Ogden, and Holzapfel (GOH) formulation to incorporate WM anisotropy. This approach was described in detail in our previous work[10]. Moreover, the fibre dispersion parameter in the GOH model was informed by FA, following the method described by Giordano and Klevin [48]. The material properties used in this study are provided in Table 1.

**Table 1.** Material coefficients for brain materials and correlation of fractional anisotropy with corresponding dispersion orientation parameter applied in the finite element head model.

| A) Material properties |  |  |  |
| --- | --- | --- | --- |
| Tissue Type | Material Description | Density | Reference |
| Skull | Rigid Body | Density: 2 kg/dm<sup>3</sup> | Kleiven(2010)[49] |
| Dura mater | Isotropic Elastic | Density: 1.13 kg/dm<sup>3</sup><br>Young Modulus: 31.5 MPa<br>Poisson's Ratio: 0.45 | Kleiven(2010)[49] |
| Falx and Tentorium | Isotropic Elastic | Density: 1.13 kg/dm<sup>3</sup><br>Young Modulus: 31.5 MPa<br>Poisson's Ratio: 0.45 | Kleiven(2010)[49] |
| CSF | Incompressible Neo-Hookean | Density: 1 kg/dm<sup>3</sup><br>Bulk Modulus: 2.1 GPa | Giordano et al. (2014)[50] |
| Gray and White Matter | Uncoupled Viscoelastic | Density: 1.04 kg/dm<sup>3</sup><br>Bulk Modulus: 50 MPa | Giordano et al. (2014)[50] |
| B) Fractional anisotropy correlation to corresponding dispersion orientation parameter[50] |  |  |  |
| Fractional anisotropy |  | k value |  |
| 0.0 – 0.2 |  | 0.3333 |  |
| 0.2 – 0.3 |  | 0.2732 |  |
| 0.3 – 0.4 |  | 0.2500 |  |
| 0.4 – 0.5 |  | 0.2273 |  |
| 0.5 – 0.6 |  | 0.2000 |  |
| 0.6 – 0.7 |  | 0.1667 |  |
| 0.7 – 0.8 |  | 0.1282 |  |
| 0.8 – 0.9 |  | 0.0769 |  |
| 0.9 – 1.0 |  | 0.0000 |  |
| Note: Gray matter regions are assigned k=0.333. Correspondence above is applicable for white matter regions. |  |  |  |

Considering the influence of WM orientation on the pattern of deformation along major fibre direction, fibre orientation was incorporated into the model following our previous approach[10,21]. Fibre direction vectors derived from the template FOD were mapped onto the FE mesh, assigning a local fibre orientation to each WM element. These element-wise fibre vectors were then used to define the preferred fibre direction in the fibre-reinforced hyper-viscoelastic constitutive model implemented in the open-source FE software called FEBio (www.febio.org). The final FE model has edge length ranging from 1.5 mm to 5.0 mm, and 92,736 elements.

To create a realistic representation of the brain-skull interface, CSF and dura layers are included in the model. The contact interface between the brain and the skull was modelled using a sliding elastic with friction factor of 0.2[51]. The CSF elements in contact with the falx and tentorium were re-assigned as falx and tentorium and were modelled to be in tied contact with the dura layer to represent the continuity of this membrane. An overview of the full pipeline is presented in Figure 1.

**Figure 1.**
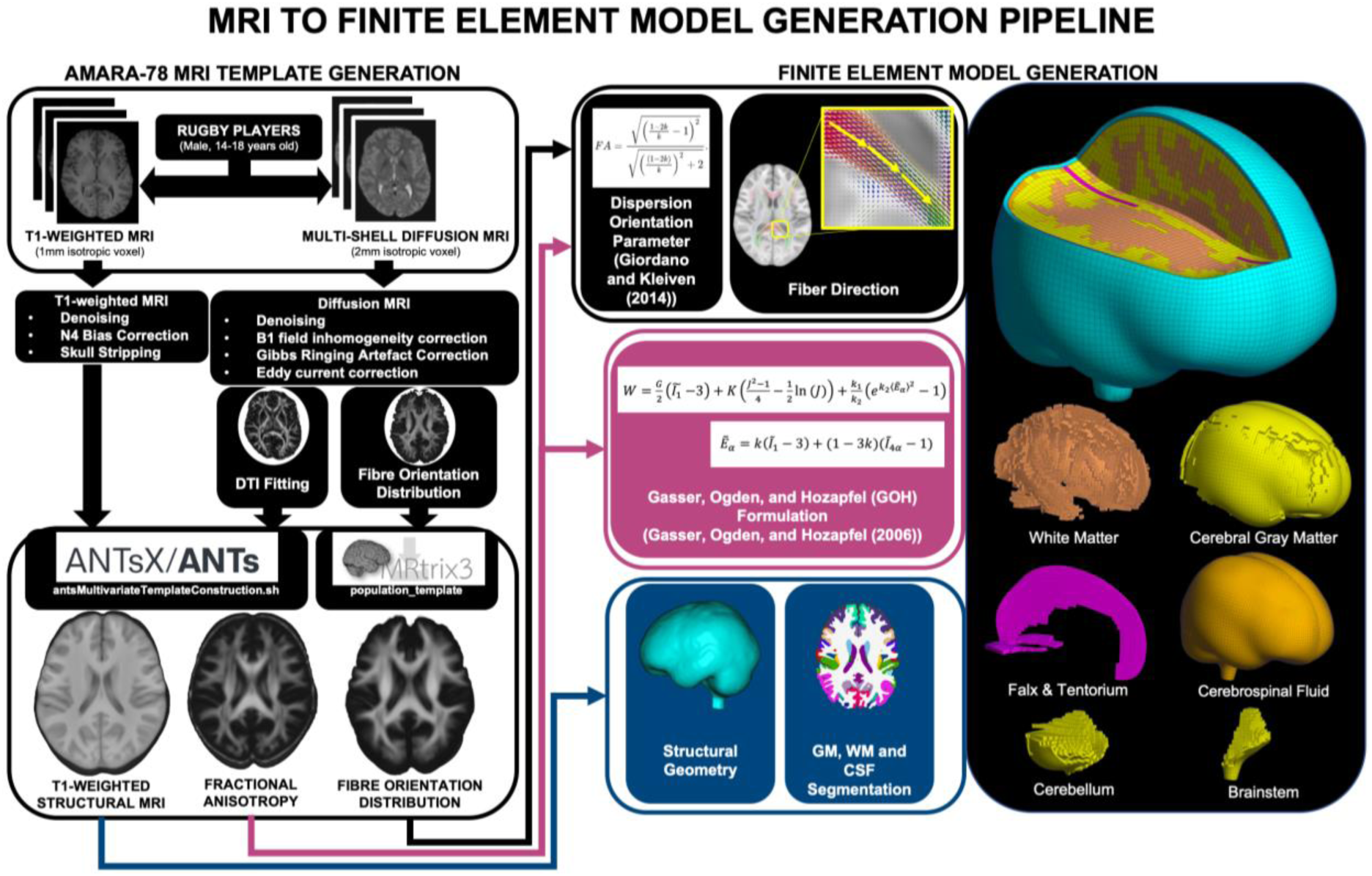
**Summary figure illustrating the pipeline from MRI processing to finite element head model generation.**

### FE model validation

After the FE model was created, validation is required to assess the biofidelity of the results. In this work, validation is performed against the experiments of Alshareef et al. and Hardy et al. on cadaveric head impacts[52,53].

Alshareef et al. applied pure coronal rotation on the cadaveric head (male, 53 years old) using a rotational test device. Subsequently, the displacement results for crystals in the skull were recorded. To replicate this loading condition in FEBio, a prescribed rotational motion was applied to the model using a load curve that peaks at 20 rad/s over 60 ms. The rotation was applied about the coronal (x) axis, producing side-to-side rotation of the head. The locations of the experimental receivers (receivers 9, 16, and 31) were approximated based on the reported x-, y-, and z-coordinates relative to the model’s centre of gravity, identified using the same process described by Alshareef et al. (*i.e.*, use of Frankfort plane). The corresponding brain tissue nodes were identified, and triaxial displacements were extracted relative to their initial positions for comparison with the experimental measurements.

Similarly, the tri-axial linear and rotational acceleration reported by Hardy et al. was also applied to the model. For this validation, C064-T2 (male, 82 years old) was used, and neutral density targets (NDT) 7 and 8 were approximated in the FE head model based on the provided initial position. Displacement of these NDTs with respect to the centre of gravity is recorded and compared with those reported in the literature. This test was chosen as a reference to reduce effect of sex on validation as well as it being sufficiently reported in the supplementary material of Hardy et al.(2007).

CORrelation and Analysis (CORA) score was computed by incorporating both corridor rating and cross-correlation rating as implemented in the CORA tool by Gehre et al. [54]. The corridor method evaluates the deviation of the simulated response from experimental data within automatically defined corridors, whereas the cross-correlation method quantifies agreement in phase, size, and progression. The final CORA score is calculated as a weighted sum of the corridor and cross-correlation ratings.

### Subject-specific model generation from the AMARA-78 MRI template

After the template model validation, the ability of the pipeline to create subject-specific models was evaluated. From our cohort, one athlete exposed to repeated HAE throughout the season was randomly selected. A subject-specific brain surface mesh was generated from the participant’s T1-weighted brain mask and used as the target geometry for morphing the template FE model.

To match the subject’s unique brain shape, the free-form deformation algorithm [55] developed in-house was applied to morph the template FE mesh model into the subject’s surface mesh. The target surface mesh contained substantially more vertices than the external nodes of the template FE model, providing a sufficiently dense set of correspondence points to achieve a stable root mean square error between template and target geometry (<1mm). This morphing method preserves the nodal and elemental relationships of the template mesh across all subject-specific models, allowing direct inter-subject comparison in future large-cohort studies. After mesh morphing, element-wise assignments, including material properties, anatomical label, and fibre direction, were performed in the same manner as for the template FE model, and contact interfaces were similarly defined.

Instrumented mouthguards data from all recorded HAE across the entire cohort for the whole season were pooled to derive the 95^th^and 99^th^ percentile highest-intensity HAE based on peak angular acceleration representing high-severity exposure within our sample. These loading conditions were subsequently applied as boundary conditions. This approach provides determination of upper-bound representation of in-practice and in-game HAEs recorded during the season, enabling model comparison without relying on single-subject extreme events that may be influenced by subject-specific variability. Details of this HAE and its profile can be accessed in Supplementary Material 2. Briefly, the 95th percentile impact had a peak angular acceleration of 3,416 rad/s², whereas the 99th percentile impact reached 6,371 rad/s². For this analysis, three models were compared: 1) the template model with WM and GM assigned as isotropic materials (k=0.333), 2) the template model incorporating a dispersion orientation parameter for heterogeneous WM materials, and 3) a fully subject-specific model generated from the subject’s MRI. After simulation, 1^st^ principal Green-Lagrange strain (maximum principal strain – MPS) was extracted and averaged within each region using the Desikan-Killiany[42] parcellation. Regional averaging was used to summarize the spatially varying strain field into an anatomically meaningful regional measures, enabling robust comparison of strain patterns across models within a standardized neuroanatomical framework.

Regional averaged strains were extracted for each model at 10, 20, 30, 40, and 50 milliseconds. Considering the nested structure of the data – 84 regions evaluated across five discrete time points – a linear mixed-effects model (LMM) was used to evaluate the influence of model type on brain response. This accounted for the non-independence of spatial measurements and the repeated-measures nature of strain profiles. Average maximum principal strain served as the dependent variable, while model type and timestep were defined as fixed effects. Region of interest was incorporated as a random effect. To address multiple comparisons, p-values were adjusted using the Benjamini-Hochberg[56] procedure to control the false discovery rate (FDR).

## RESULTS

### AMARA-78 TEMPLATE EVALUATION

The AMARA-78 T1-weighted and FA template generated from rugby players are illustrated in Figure 2 which shows representative axial slices of the T1-weighted template and the corresponding FA template. The volume of the template cerebellar and cerebral cortex GM and WM, including comparison with the input rugby players are presented in Supplementary Material 3.

**Figure 2.**
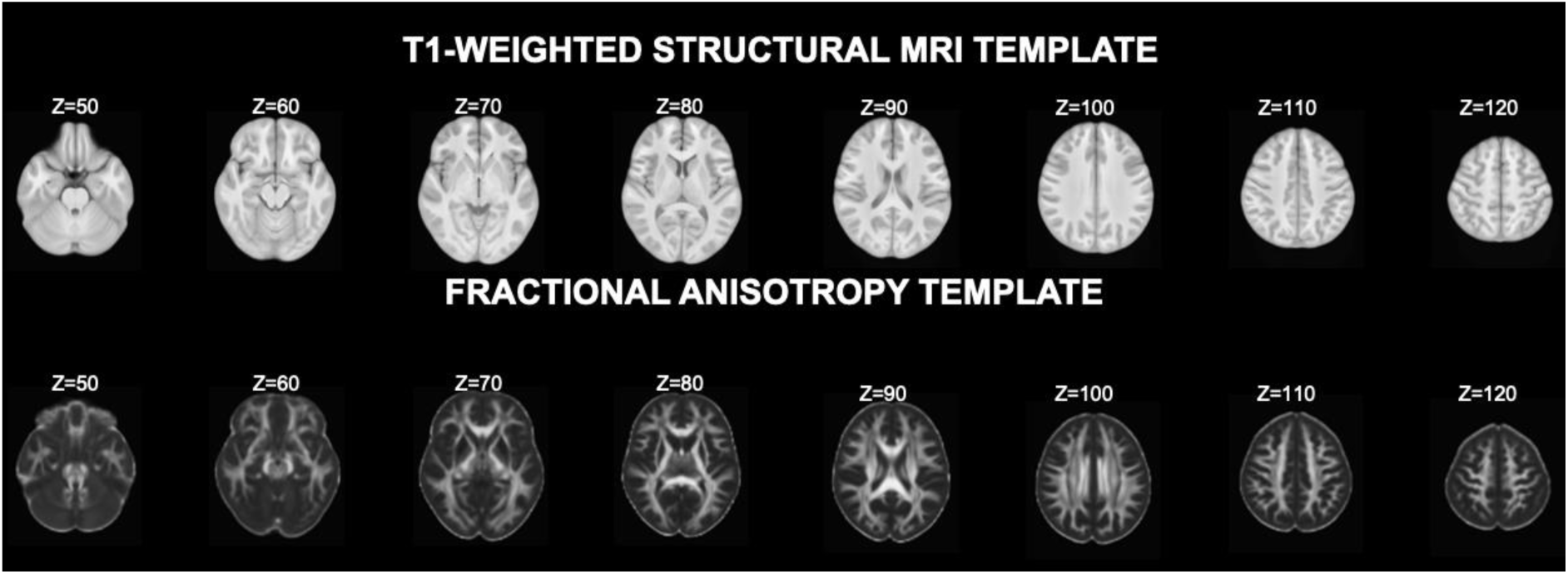
**Representative axial slices of the T1-weighted (top) and the corresponding fractional anisotropy template (bottom).**

After generating the template, we evaluated whether AMARA-78 provided a better anatomical reference for this cohort than existing reference templates. This was assessed using deformation-based morphometry done on three templates – AMARA-78, MNI152 and PNG. The amount of local anatomical warping required during registration was quantified using the absolute log-Jacobian determinant |logJ|. Larger |logJ| values indicate greater local expansion or compression, and therefore greater anatomical deformation, during normalisation. As shown in Figure 3, fewer voxels showed significantly larger |logJ| when using the AMARA-78 than MNI152-1mm, (MNI152-1mm: 140,148 voxels; AMARA-78: 94,920 voxels). Similarly, registration to AMARA-78 revealed fewer voxels showing significant larger |logJ| compared to PNG (PNG: 376,624 voxels; AMARA-78: 169,762 voxels).

**Figure 3.**
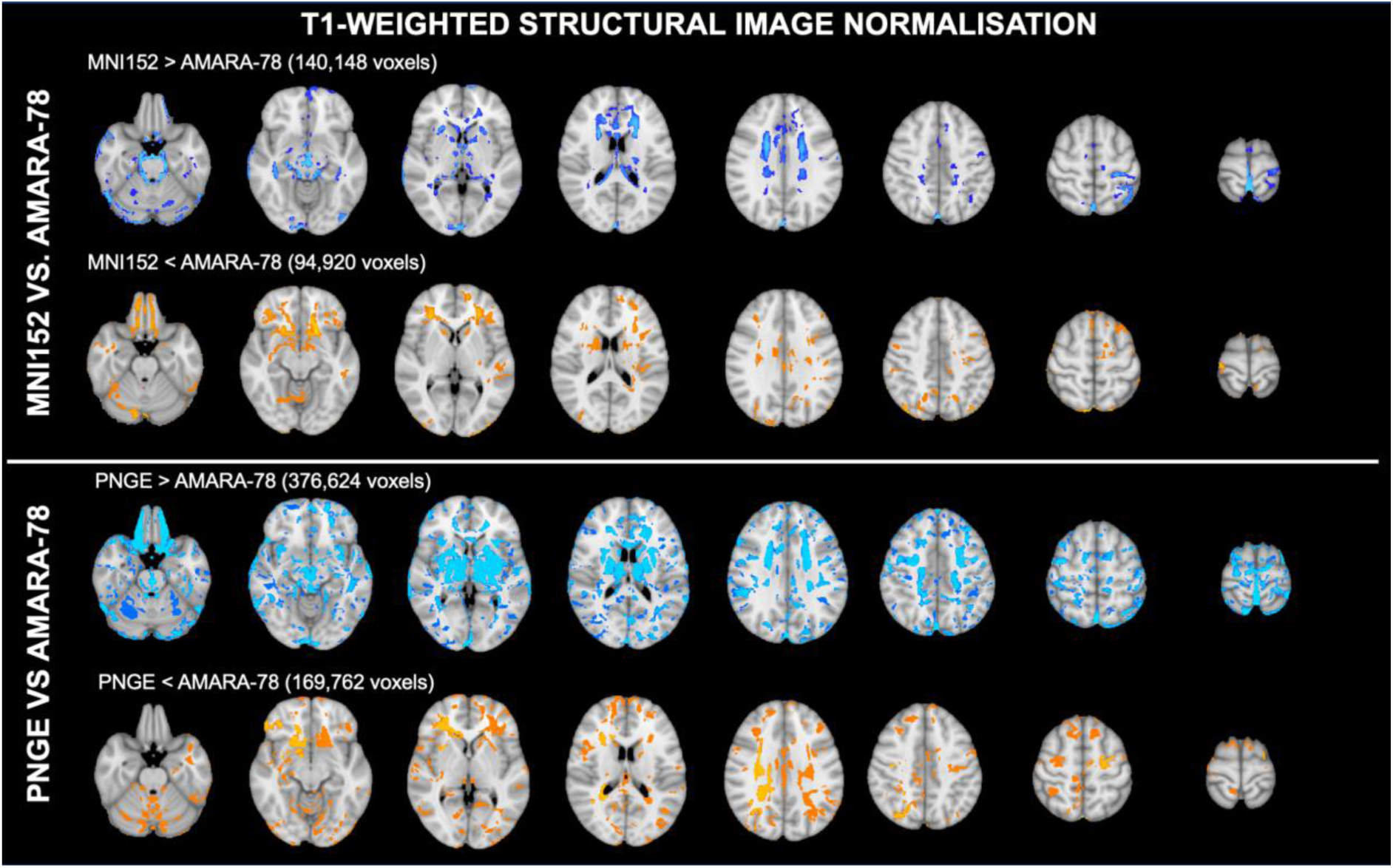
Voxel-wise statistical maps (p < 0.05, FWE-corrected) of deformation (|logJ|) in non-collision sport controls, comparing normalisation to the MNI152-1 mm and PNG template with normalisation to AMARA-78. Results are shown in the axial plane. Red and blue regions indicate significantly greater and lesser morphological change, respectively, during spatial normalisation relative to the proposed template.

## FE MODEL VALIDATION

The FE head model developed from the AMARA template was validated against two sets of experimental data using CORA to quantify the difference between the FE head model-predicted displacements and those recorded in the experimental data. CORA scores were qualitatively interpreted following the effect size convention according to this scale[57]: small (<0.2), moderate(0.21-0.50), and large (≥0.8).

### Validation against Alshareef et al

The displacement responses along the x-, y-, and z-axes were extracted from FE brain nodes corresponding to experimental receivers 9, 16, and 31 reported by Alshareef et al. These responses are compared with experimental measurements in Figure 4. Quantitative agreement was assessed using CORA scores, with results for each receiver–axis pair summarised in Table 3. Overall, CORA analysis indicated moderate agreement between simulated and experimental responses, with scores ranging from 0.30 to 0.36.

**Figure 4.**
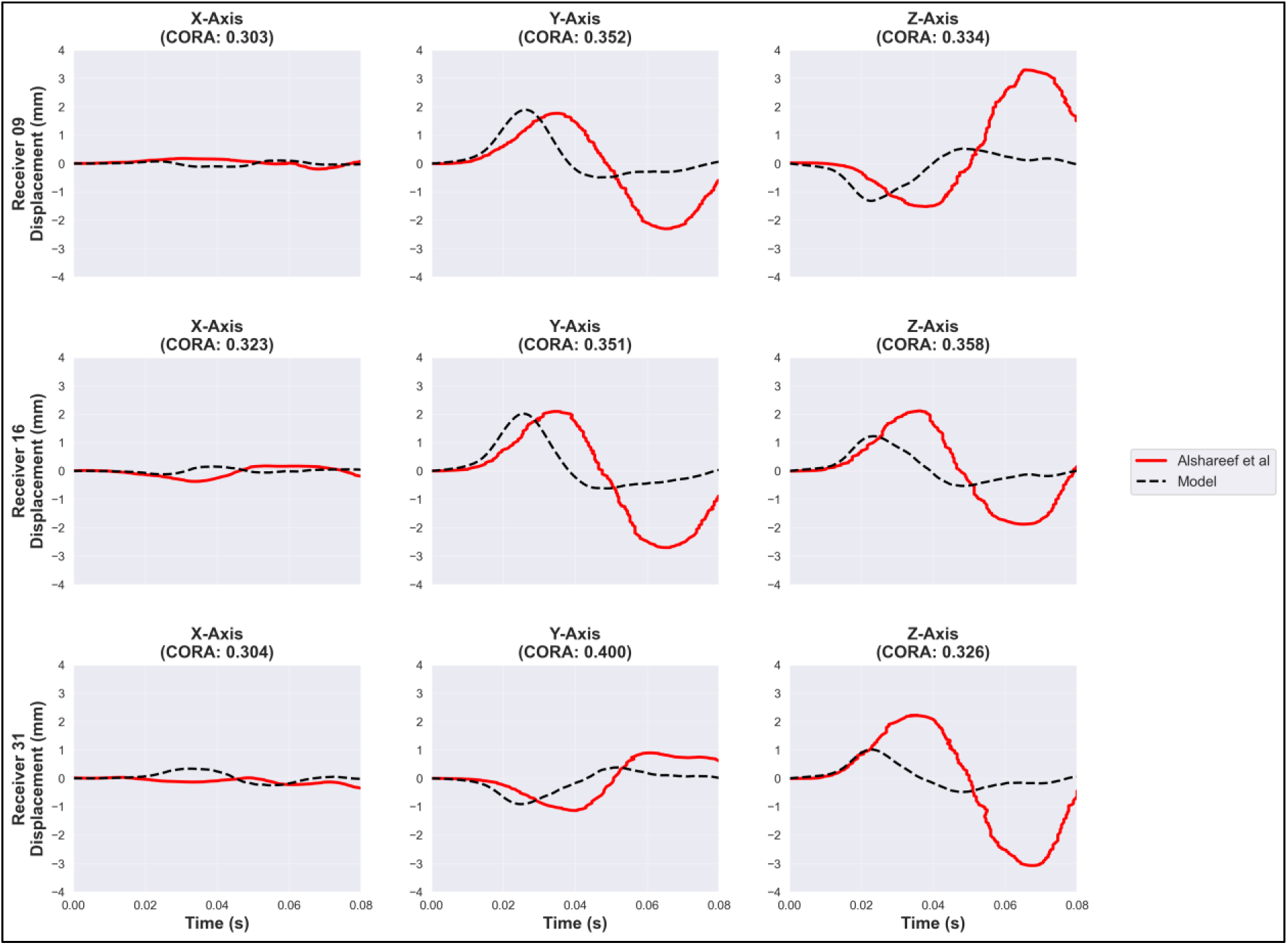
**Comparison between experimental data from Alshareef et al. and model simulation**

**Table 3.** CORA scores of observed nodal displacements in the finite element head model compared versus cadaveric head experiments.

|  |  |  |  |
| --- | --- | --- | --- |
| Experiment: Alshareef et al. |  |  |  |
| Receiver | 9 | 16 | 31 |
| Axis -X | 0.30 | 0.32 | 0.30 |
| Axis -Y | 0.35 | 0.35 | 0.40 |
| Axis -Z | 0.33 | 0.36 | 0.33 |
| Experiment: Hardy et al. |  |  |  |
| NDT | 7 | 8 | -- |
| Axis -X | 0.48 | 0.65 | -- |
| Axis -Z | 0.57 | 0.60 | -- |

**Table 4.** Time-series mean and standard deviation of the average maximum principal strain across all regions of interest, as defined by the Desikan–Killiany atlas, for the isotropic template model, the template model incorporating a dispersion orientation parameter, and the subject-specific finite element head model.

| Timestep (ms) | Isotropic Template Model | Template Model with Dispersion Orientation Parameter | Subject-specific |
| --- | --- | --- | --- |
| **95<sup>TH</sup> PERCENTILE IMPACT** |  |  |  |
| **10** | 0.018 [0.016-0.021] | 0.022 [0.018-0.027] | 0.025 [0.021-0.029] |
| **20** | 0.033 [0.023-0.040] | 0.037 [0.027-0.045] | 0.046 [0.035-0.058] |
| **30** | 0.017 [0.015-0.020] | 0.026 [0.021-0.032] | 0.033 [0.027-0.039] |
| **40** | 0.008 [0.007-0.009] | 0.017 [0.014-0.019] | 0.013 [0.012-0.016] |
| **99<sup>TH</sup> PERCENTILE IMPACT** |  |  |  |
| **10** | 0.021 [0.017-0.026] | 0.021 [0.017-0.026] | 0.026 [0.021-0.030] |
| **20** | 0.076 [0.061-0.091] | 0.077 [0.061-0.092] | 0.071 [0.054-0.090] |
| **30** | 0.098 [0.083-0.117] | 0.096 [0.082-0.115] | 0.127 [0.105-0.154] |
| **40** | 0.036 [0.031-0.044] | 0.035 [0.031-0.043] | 0.035 [0.030-0.040] |

### Validation against Hardy et al

Similarly, displacement responses along all three axes were obtained from nodes corresponding to neutral density targets (NDTs) 7 and 8 from Hardy et al.’s experiments. Comparisons between simulated and experimental data are shown in Figure 5. CORA scores for each NDT–axis combination are reported in Table 3. Overall, CORA analysis indicated moderate scores ranging from 0.48 to 0.65.

**Figure 5.**
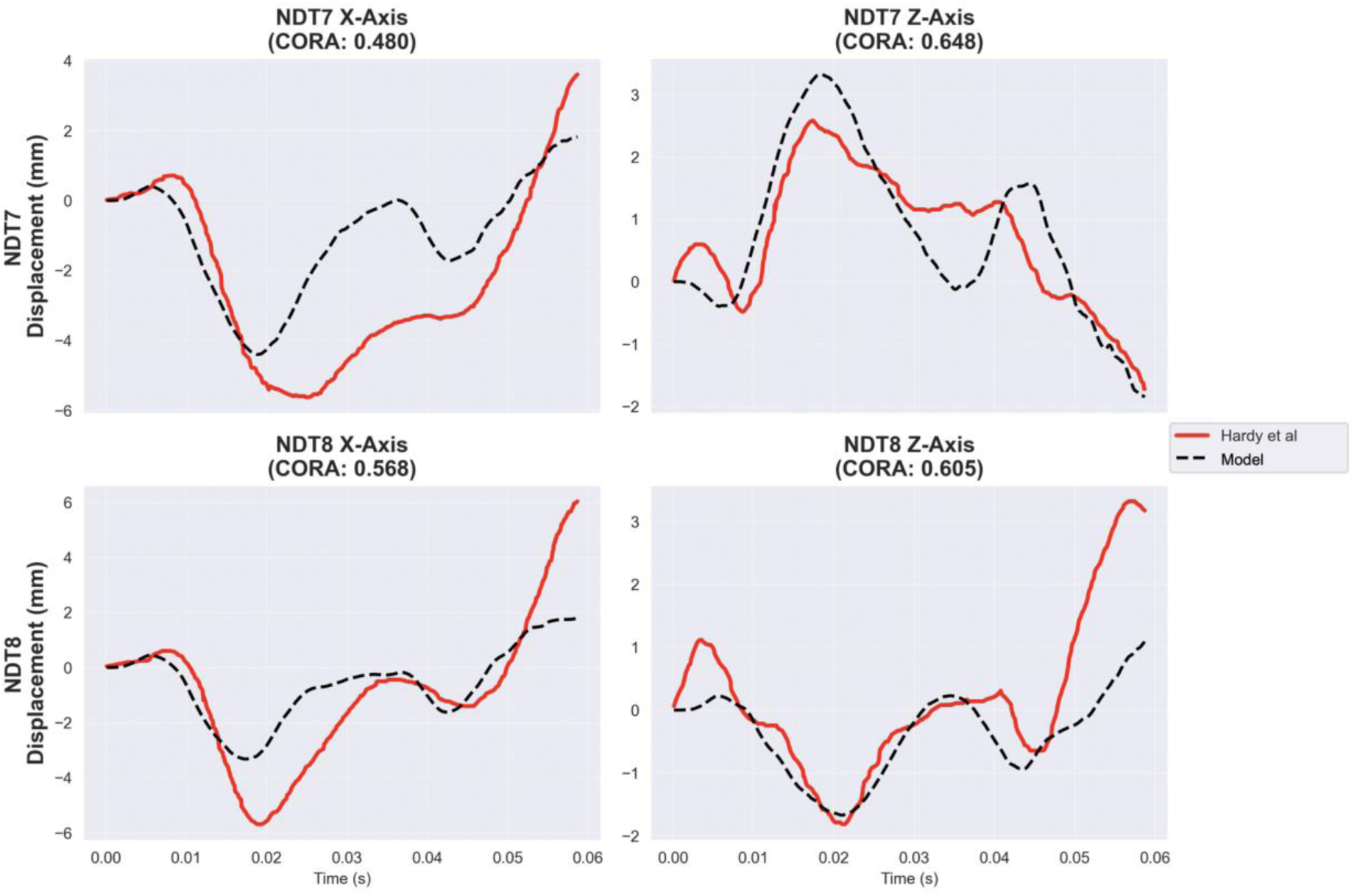
**Comparison between experimental data from Hardy et al. and model simulation**

## COMPARISON OF TEMPLATE AND SUBJECT-SPECIFIC FE HEAD MODEL

After validating the AMARA-78 FE head model against experimental displacement data, we evaluated how different levels of model personalisation influenced predicted brain strain. Three model configurations were compared– 1) isotropic template model, 2) the template model incorporating spatially varying FA-derived fiber dispersions and orientation parameters, and 3) a fully subject-specific FE head model, This comparison was performed to determine whether incorporating MRI-derived white matter information into the template model altered strain predictions relative to an isotropic template baseline, and how these template-based predictions compared with a fully subject-specific model.

Before comparing strain predictions, the MRI-to-FE assignment of diffusion-derived material information was visually assessed. Figure 6 compares the original FA map with the FE model after element-wise FA assignment. This qualitative comparison demonstrates the spatial distribution of FA values was preserved when voxel-based MRI information was transferred to the FE mesh. The template FA map appears smoother or less locally heterogeneous than subject-specific FA map because it represents a population average.

**Figure 6.**
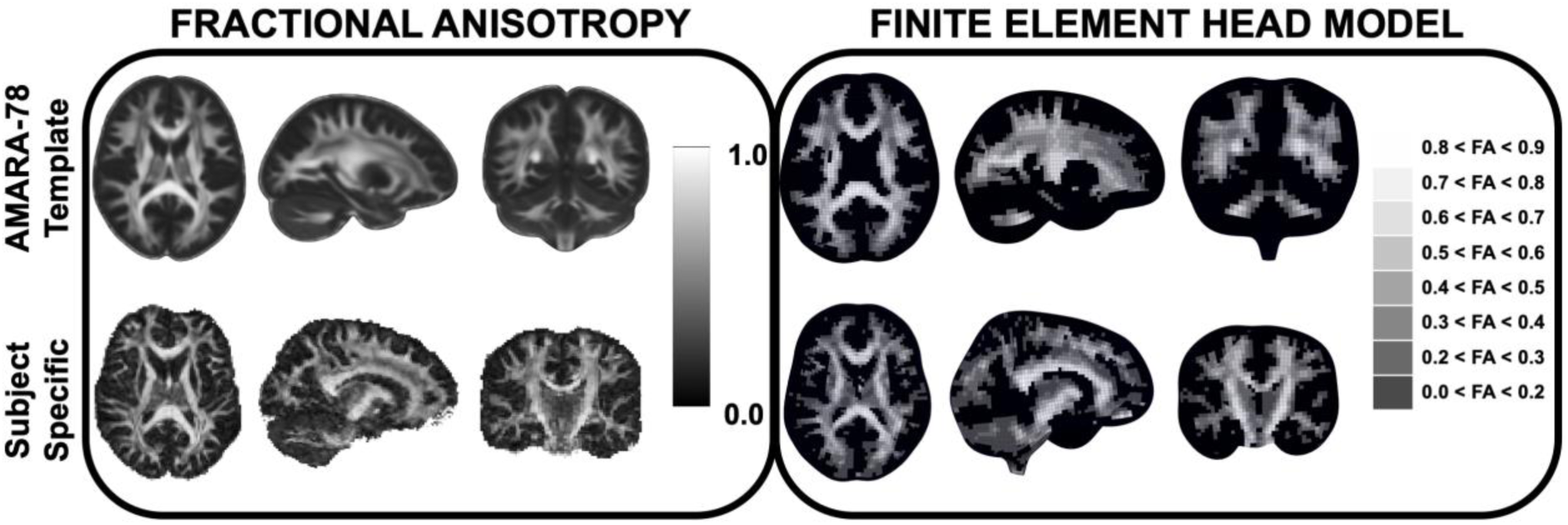
Qualitative comparison of the fractional anisotropy map and the finite element head model, colour-coded based on the fractional anisotropy value. The template FA map appears smoother or less locally heterogeneous than the subject-specific FA map because it represents a population average. When FA values are mapped from MRI to the FE model, this same spatial pattern is retained in the corresponding FE model

These models were then subjected to the 95^th^ and 99^th^ percentile impact as boundary conditions, and the resulting maximum principal strain was compared across atlas-defined brain regions as shown in Figure 7 and 8. Generally, the regional strains across all models peaked between 20 ms and 30 ms before decreasing at 40 ms. While the three models yielded varying strain magnitudes under the 95th percentile impact, the differences between the isotropic template and the template model with dispersion diminished at the 99th percentile impact; in contrast, the subject-specific model maintained a distinct profile across both impact intensities.

**Figure 7.**
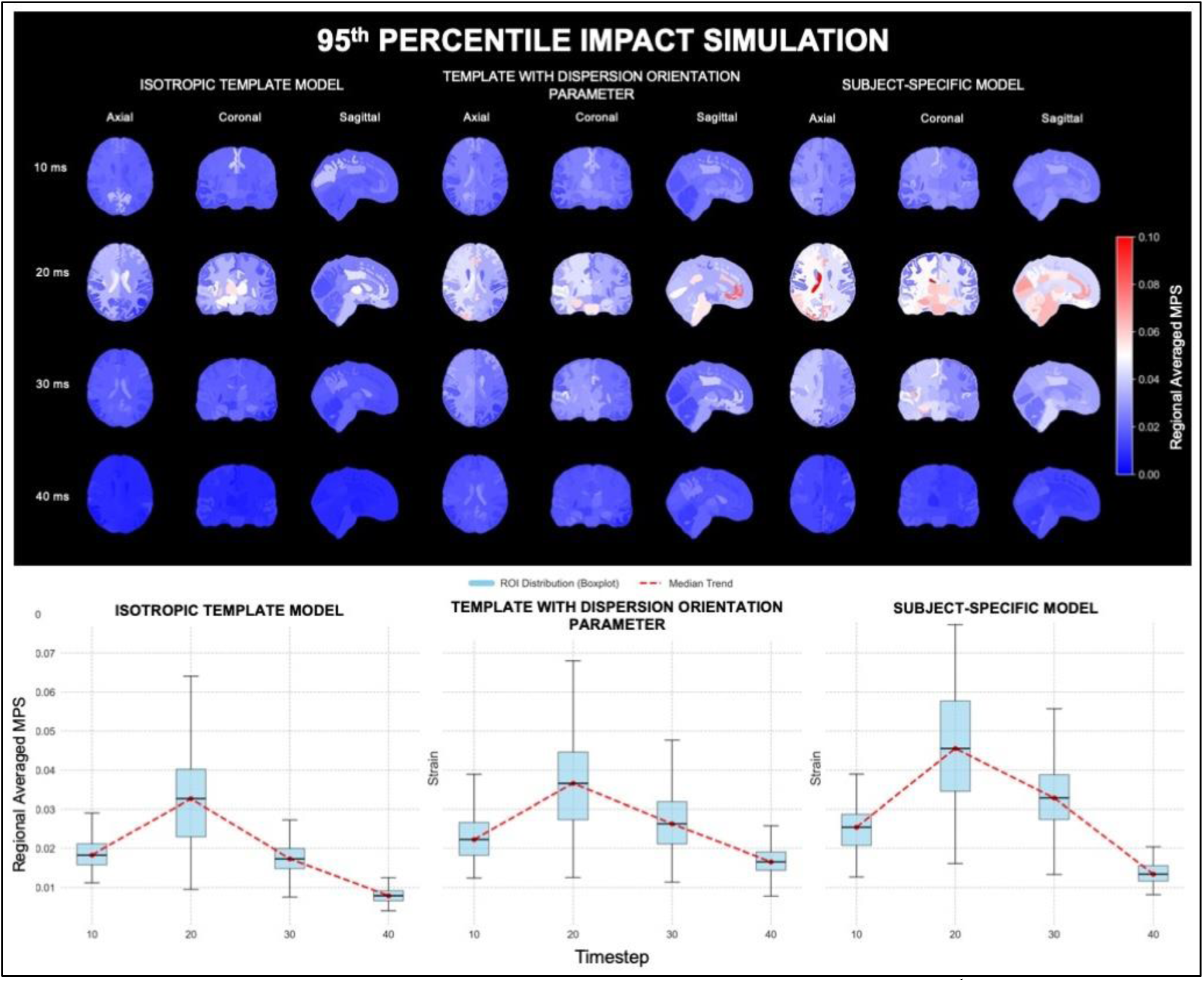
A) Spatial maximum principal strain for the whole brain for the three models under the 95^th^ percentile impact based on peak angular acceleration across the whole rugby season for all athletes as a boundary condition. B) Time-series of average maximum principal strain across regions of interest defined by the Desikan–Killiany atlas, comparing the isotropic template, the template incorporating a dispersion orientation parameter, and the subject-specific model.

**Figure 8.**
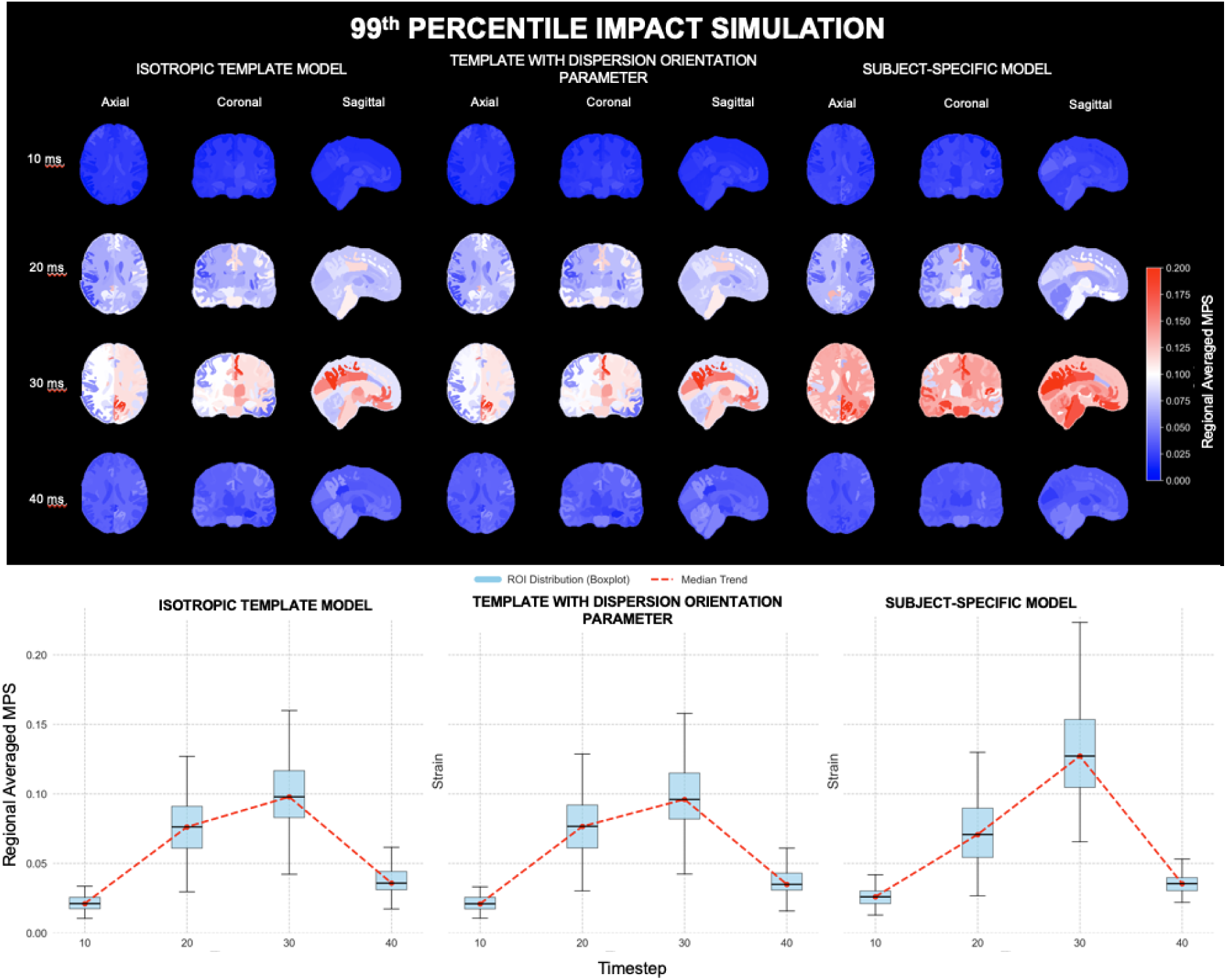
A) Spatial maximum principal strain for the whole brain for the three models under the 99^th^ percentile impact based on peak angular acceleration across the whole rugby season for all athletes as a boundary condition. B) Time-series of average maximum principal strain across regions of interest defined by the Desikan–Killiany atlas, comparing the isotropic template, the template incorporating a dispersion orientation parameter, and the subject-specific model.

The LMM demonstrated that model construction significantly influenced predicted maximum principal strain across the brain (p < 0.01). At the 95th percentile impact condition, both the template model incorporating fibre dispersion parameters and the subject-specific model showed significant differences compared with the isotropic template baseline (p < 0.01), In contrast, at the 99th percentile impact condition, only the subject-specific model exhibited a significant difference relative to the isotropic template, while the template model with fibre dispersion did not. These differences remained statistically significant after FDR correction, indicating a robust effect of model construction on strain prediction. Timestep also significantly influenced strain magnitude (p < 0.01), consistent with the observed time-dependent strain response across models.

## DISCUSSION

In this study, a computational framework for the generation of a population-specific brain template was established. Subsequently, this pipeline was extended to create an FE head model with heterogeneous material properties guided by dMRI-derived metrics, further increasing its bio-fidelity. This was validated against human cadaveric head experiments conducted by Alshareef et al.[52] and Hardy et al.[53], demonstrating that this pipeline is able to capture the transient biomechanical response of the head under HAE. Finally, a comparison of the strain profile in the brain upon exposure to the 95^th^ and 99 ^th^ percentile HAE recorded across the season was made between the isotropic model, MRI material-guided template model, and subject-specific models.

### Average template generation

HAE exposure and mTBI diagnosis are more common in athletes participating in collision sports than in non-contact sports[58]. Potentially, due to repetitive HAE exposure, the brains of these athletes may exhibit a distinct trajectory from non-contact sport or age-matched counterparts[9]. Emerging evidence suggests that repetitive HAE exposure may alter neuroanatomical trajectories, as evidenced by dMRI studies probing microstructural divergence in athlete cohorts [59–61]. Despite these findings, the literature remains characterised by heterogeneous results, often attributed to variability in MRI processing pipelines [60]. A critical but frequently overlooked factor in this heterogeneity is the systematic bias introduced during spatial normalisation.

In a study of Wilke et al.[62], a strong age effect was found when comparing the normalisation of paediatric subjects to a standard adult and a custom paediatric template. In the present study, we observed a greater volume of voxels exhibiting substantial deformation when registering our control cohort to the MNI152 atlas than when registering it to our AMARA-78 template. This difference is likely influenced, at least in part, by the age discrepancy between the subjects used to construct each template. The MNI152 was generated from a cohort of adults aged 18-44 years old[63]. Furthermore, this age-related differences is further investigated by comparing normalisation to our AMARA-78 template and the PNG which are from considerably similar adolescent cohorts. Despite the similarity in age, the PNG template still showed a greater volume of voxels with larger deformations than the AMARA-78 template. This finding suggests that, although age remains an important consideration due to ongoing neurodevelopment during adolescence, other factors such as demographic differences may also contribute to morphological variability leading to template-specific deformation patterns [64]. While this does not inherently invalidate MNI152 as a standard space for general populations and the utilization of PNG for adolescent cohorts, it highlights the critical need for age-and cohort-appropriate templates in sports-related neurotrauma research. Moreso, this adds up to the increasing literature highlighting the necessity to use study cohort specific template not only for adolescents, but also for pathological or atrophy related studies [65].

A difference in the underlying anatomy of a study cohort from the target template can lead to mis-registration, which can introduce bias, leading to errors in the analysis. Zou et al. demonstrated that using a population-specific diffusion MRI template for early-to-middle adolescent collision-sport athletes reduced spatial normalisation bias and improved the sensitivity of for detecting regional white matter diffusion changes compared with existing standard templates[9]. By utilising a template that reflects the specific morphometry of our rugby cohort, we minimise registration-induced bias, helping ensure that observed biomechanical and structural variances are reflect true physiological differences rather than artefacts of the normalisation process.

### Finite element head model

This study integrates brain MRI with head impact FE simulations by generating an FE head model from the MRI-derived population template. FE analysis of the mechanical response of the brain under HAE has become widely used in TBI research [21], primarily because it provides regionalised strain profile that allow further characterisation of potential alterations caused by HAE. Miller et al., in a study relating HAE metrics to longitudinal DTI changes in youth football players, showed that strain-based cumulative exposure metrics were better predictors of WM diffusion changes than traditional kinematic-based impact metrics[66]. However, while dozens of FE models have been developed in the past[67], their bio-fidelic property to mimic real world brain response is of utmost importance. A gold standard for validation is to compare FE simulation results with cadaveric head experiments[52,53]. Given that the proposed MRI-to-FE pipeline in this study was designed for TBI, mTBI, and/or HAE studies, brain deformation is a relevant mechanism to consider[68]. Therefore, the model generated through the pipeline was validated against cadaveric head experiments.

Validation of our FE model revealed small to moderate CORA score, which are comparable to those reported FE head model studies that performed similar validations [5,20]. As discussed in depth by Carmo et al., these differences can be explained by several factors [20], primarily, in this case, age difference. The MRI template used to generate the FE model was from adolescent rugby players aged 14-18 years, whereas Alshareef et. al. used a 53-year-old specimen, and Hardy et al. used an 82-year-old male. Kleiven et al. have shown the effects of brain size and shape on the brain’s mechanical response to impact[6]. Nevertheless, qualitatively, the behaviour of our FE model, measured by the displacement magnitude (*i.e.,* 0-3 mm for Alshareef et al.) of the corresponding nodes relative to the receivers or NDTs used in the cadaveric experiments, closely matched the experimental data. While differences in size and geometry between our model and the cadaveric head specimens may explain the deviation in brain response between our FE model and the cadaveric experiments, they also highlight the importance of the pipeline constructed in this study, which includes generating a population-appropriate template and FE model.

This pipeline validation provides confidence in biofidelity and the concordance of strain measures from simulation with real-life brain response. This can provide insights into the biomechanics of brain injury and deformation by warranting estimation of regional strain from measured head kinematics. Moreover, given that this pipeline uses open-access packages and software[30,32,69,70], other studies can replicate and apply it to their study cohort without being limited by access to proprietary algorithms. This may help address the lack of sufficient data across different sports, sex, and age groups, as emphasised by Ji et al.[68].

Another finding of this study was that strain patterns differed between the template models with and without the dispersion orientation parameter and the subject-specific model at low-intensity impacts, but these differences diminished at high-intensity impact. While qualitative, the divergence observed at lower intensity impact underscores the importance of constitutive complexity. In these cases, brain material properties may act as a primary modulator of deformation.

Although the influence of brain material differences in anatomically detailed head models remains to be fully investigated [71], model features such as geometry, material properties and fibre orientation can play an important role in predicting brain deformation in a continuous and non-linear manner [72]. Across loading conditions ranging from subconcussive to mTBI level impacts, increasing impact severity is generally expected to produce larger mechanical responses This continuity assumption is reasonable across a broad biomechanical spectrum of HAE, but may become invalid at higher impact severities due to mechanical breakdown. It should be noted that, under high-intensity impact, only the subject-specific model showed a significant difference compared with the two template models. This difference is potentially driven more by intracranial volume differences than by variation in material properties. Kleiven and von Holst have shown that scaling a 3D adult FE head model across six varying brain sizes resulted in increasing brain response[6].

A study highlighted variation in the validation scores of different brain material models when validated against cadaveric head experiments and low-intensity volunteer tests [72]. Specifically, some constitutive formulations achieved high CORA scores when validated against cadaveric experiments but performed less favourably under low-intensity volunteer tests, highlighting the sensitivity of model performance to both material properties and loading conditions. Similarly, the difference in CORA scores observed between validation against the pure coronal rotation experiments of Alshareef et al.[52] and 6-degree-of-freedom experiments of Hardy et al.[53] may reflect not only differences in impact severity, but also differences in kinematic loading. In addition, while both experiments involved comparable peak angular velocities (∼20 rad/s), the peak angular accelerations differed substantially, at approximately 1,000 rad/s² in Alshareef et al. and 4,000 rad/s² in Hardy et al. Although optimization of the material properties of the FE model was beyond the primary objective of this study, future work should consider validating the reliability of FE model under varying boundary conditions, including both low-and high-intensity impacts.

A novel contribution of this work is our MRI-to-FE pipeline, especially the application of our element assignment algorithm [73], allowing direct correspondence between FE elements and voxels in the MRI image. Initial application of this has been applied to our template model and has also been shown to be applicable for generating subject-specific FE head models. Therefore, rather than using separate model parts to represent different brain regions with distinct material properties, this approach allows a unified representation of spatially varying material properties within a single brain model. To account for differences in brain size and shape, our pipeline uses free-form deformation, which has previously been demonstrated not only for the brain, but for multiple other organs [10,47,55].

Furthermore, the incorporation of a dispersion orientation parameter better accounts for the effect of anisotropy on the brain’s mechanical response. In this study, comparison of the isotropic and anisotropic FE head models generated from the template MRI showed that maximum principal strain varied depending on model complexity. The higher strain observed in models incorporating the dispersion orientation parameter agrees with Giordano et al.[50], who reported higher Green-Lagrange strains in anisotropic simulations compared with isotropic counterparts. This further highlights the importance of incorporating anisotropic material parameters in FE head models. While subject-specificity in this study pertains to geometry and dispersion orientation parameter, previous studies have explored metrics derived from magnetic resonance elastography and tagged-MRI, which may be incorporated either to improve model biofidelity or provide additional validation[68,74–77]. Future investigations incorporating these measures would benefit from our element assignment algorithm, enabling efficient assignment of MRI-derived material properties to the FE model.

### Limitation

Limitations of this study include the use of a sex-specific cohort, which limits the ability to assess whether template generation also resolves sex-specific differences in brain response. However, this study sought to develop a pipeline that can be applied to different cohorts, defined by age, sex, ethnicity, or other population characteristics. Future investigation will include the collection of data from female subjects.

In addition, no additional event-level verification, thresholding or filtering of the recorded iMG data was performed beyond the manufacture’s processing pipelines. These iMGs have been validated by manufacturers and other research groups; however, future investigations involving large-scale FE simulations with recorded iMG data should include a dedicated processing pipeline for event verification and application of recorded HAE as boundary conditions to the model.

Finally, the investigation of how subject-specific FE model generation affects strain-profile outputs is limited. While this was performed on a single subject, significant differences in brain strain responses were observed between the isotropic template, the anisotropic template, and subject-specific models. This warrants further investigation into the added value of subject-specific FE models.

## Conclusion

In this study, we have developed a computational framework that can generate a brain atlas from MRI scans of rugby players. This atlas was then used as input to create an FE head model incorporating a fibre dispersion parameter derived from diffusion MRI. Validation against cadaveric head experiments reveals good agreement, presenting the biofidelity of the model and providing confidence in the predicted strain responses. Moreover, comparison of an isotropic template FE model, an anisotropic template FE model incorporating fibre dispersion parameter, and a subject-specific FE model revealed significant differences in predicted regional maximum principal strain, highlighting the need to incorporate cohort and subject-specific information into the FE model.

This framework may play an important role in TBI and mTBI study by enabling the generation of population-, sex-, and ethnicity-specific FE head models, as well as fully subject-specific models that better capture the brain response of study cohorts under subject-specific head loading conditions.

## SUPPLEMENTARY MATERIAL

**Supplementary Material 1.** Gray matter parcellation using SynthSeg. Cortex (ctx), Left Hemisphere(lh), Right Hemisphere (rh)

| Index | Code | Label |
| --- | --- | --- |
| 1 | L.BSTS | ctx-lh-bankssts |
| 2 | L.CACG | ctx-lh-caudalanteriorcingulate |
| 3 | L.CMFG | ctx-lh-caudalmiddlefrontal |
| 4 | L.CU | ctx-lh-cuneus |
| 5 | L.EC | ctx-lh-entorhinal |
| 6 | L.FG | ctx-lh-fusiform |
| 7 | L.IPG | ctx-lh-inferiorparietal |
| 8 | L.ITG | ctx-lh-inferiortemporal |
| 9 | L.ICG | ctx-lh-isthmuscingulate |
| 10 | L.LOG | ctx-lh-lateraloccipital |
| 11 | L.LOFG | ctx-lh-lateralorbitofrontal |
| 12 | L.LG | ctx-lh-lingual |
| 13 | L.MOFG | ctx-lh-medialorbitofrontal |
| 14 | L.MTG | ctx-lh-middletemporal |
| 15 | L.PHIG | ctx-lh-parahippocampal |
| 16 | L.PaCG | ctx-lh-paracentral |
| 17 | L.POP | ctx-lh-parsopercularis |
| 18 | L.POR | ctx-lh-parsorbitalis |
| 19 | L.PTR | ctx-lh-parstriangularis |
| 20 | L.PCAL | ctx-lh-pericalcarine |
| 21 | L.PoCG | ctx-lh-postcentral |
| 22 | L.PCG | ctx-lh-posteriorcingulate |
| 23 | L.PrCG | ctx-lh-precentral |
| 24 | L.PCU | ctx-lh-precuneus |
| 25 | L.RACG | ctx-lh-rostralanteriorcingulate |
| 26 | L.RMFG | ctx-lh-rostralmiddlefrontal |
| 27 | L.SFG | ctx-lh-superiorfrontal |
| 28 | L.SPG | ctx-lh-superiorparietal |
| 29 | L.STG | ctx-lh-superiortemporal |
| 30 | L.SMG | ctx-lh-supramarginal |
| 31 | L.FP | ctx-lh-frontalpole |
| 32 | L.TP | ctx-lh-temporalpole |
| 33 | L.TTG | ctx-lh-transversetemporal |
| 34 | L.IN | ctx-lh-insula |
| 35 | L.CER | Left-Cerebellum-Cortex |
| 36 | L.TH | Left-Thalamus |
| 37 | L.CA | Left-Caudate |
| 38 | L.PU | Left-Putamen |
| 39 | L.PA | Left-Pallidum |
| 40 | L.HI | Left-Hippocampus |
| 41 | L.AM | Left-Amygdala |
| 42 | L.AC | Left-Accumbens-area |
| 43 | R.TH | Right-Thalamus |
| 44 | R.CA | Right-Caudate |
| 45 | R.PU | Right-Putamen |
| 46 | R.PA | Right-Pallidum |
| 47 | R.HI | Right-Hippocampus |
| 48 | R.AM | Right-Amygdala |
| 49 | R.AC | Right-Accumbens-area |
| 50 | R.BSTS | ctx-rh-bankssts |
| 51 | R.CACG | ctx-rh-caudalanteriorcingulate |
| 52 | R.CMFG | ctx-rh-caudalmiddlefrontal |
| 53 | R.CU | ctx-rh-cuneus |
| 54 | R.EC | ctx-rh-entorhinal |
| 55 | R.FG | ctx-rh-fusiform |
| 56 | R.IPG | ctx-rh-inferiorparietal |
| 57 | R.ITG | ctx-rh-inferiortemporal |
| 58 | R.ICG | ctx-rh-isthmuscingulate |
| 59 | R.LOG | ctx-rh-lateraloccipital |
| 60 | R.LOFG | ctx-rh-lateralorbitofrontal |
| 61 | R.LG | ctx-rh-lingual |
| 62 | R.MOFG | ctx-rh-medialorbitofrontal |
| 63 | R.MTG | ctx-rh-middletemporal |
| 64 | R.PHIG | ctx-rh-parahippocampal |
| 65 | R.PaCG | ctx-rh-paracentral |
| 66 | R.POP | ctx-rh-parsopercularis |
| 67 | R.POR | ctx-rh-parsorbitalis |
| 68 | R.PTR | ctx-rh-parstriangularis |
| 69 | R.PCAL | ctx-rh-pericalcarine |
| 70 | R.PoCG | ctx-rh-postcentral |
| 71 | R.PCG | ctx-rh-posteriorcingulate |
| 72 | R.PrCG | ctx-rh-precentral |
| 73 | R.PCU | ctx-rh-precuneus |
| 74 | R.RACG | ctx-rh-rostralanteriorcingulate |
| 75 | R.RMFG | ctx-rh-rostralmiddlefrontal |
| 76 | R.SFG | ctx-rh-superiorfrontal |
| 77 | R.SPG | ctx-rh-superiorparietal |
| 78 | R.STG | ctx-rh-superiortemporal |
| 79 | R.SMG | ctx-rh-supramarginal |
| 80 | R.FP | ctx-rh-frontalpole |
| 81 | R.TP | ctx-rh-temporalpole |
| 82 | R.TTG | ctx-rh-transversetemporal |
| 83 | R.IN | ctx-rh-insula |
| 84 | R.CER | Right-Cerebellum-Cortex |

**Supplementary Material 2.**
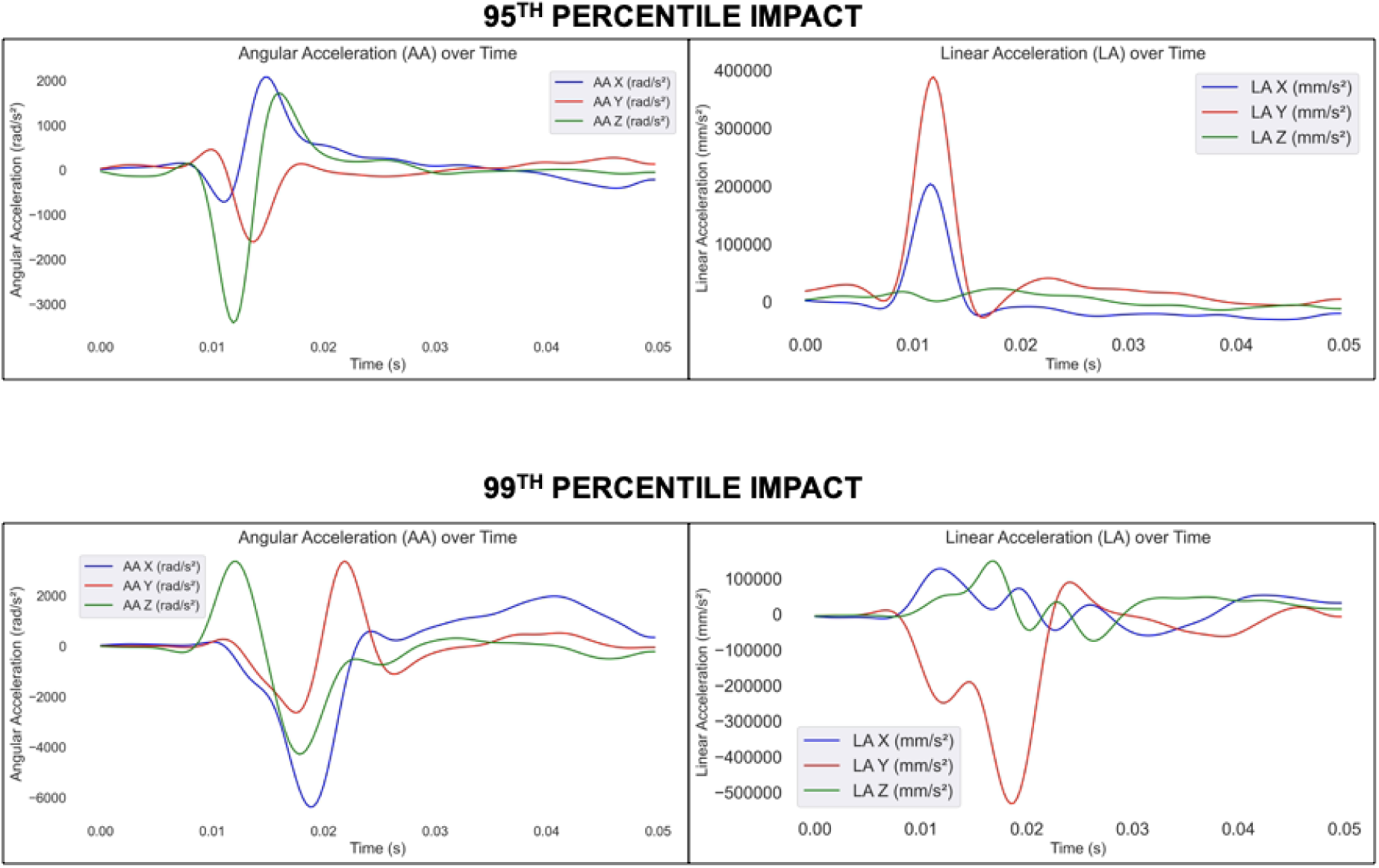
The 95^th^ and 99^th^ percentile head acceleration event used as boundary condition for comparison of isotropic template finite element head model, template with anisotropic dispersion parameter, and subject-specific model.

**Supplementary Material 3.** Table of comparison of the white matter and gray matter regional volumes between template and the mean and standard deviation of input subjects.

| STRUCTURE | Volume of Template (mm<sup>3</sup>) |  | Mean and Standard Deviation from Subjects (mm<sup>3</sup>) |  |
| --- | --- | --- | --- | --- |
| Total Intracranial | 1,557,721.10 |  | 1,594,191.62 ± 10,4675.18 |  |
| Cerebrospinal Fluid | 218,140.78 |  | 183,660.66 ± 15,486.73 |  |
| Brain Stem | 24,807.85 |  | 25,114.53 ± 2,101.52 |  |
|  | RIGHT HEMISPHERE |  | LEFT HEMISPHERE |  |
|  | Volume of Template (mm<sup>3</sup>) | Mean and Standard Deviation from Subjects (mm<sup>3</sup>) | Volume of Template (mm<sup>3</sup>) | Mean and Standard Deviation from Subjects (mm<sup>3</sup>) |
| Cerebral White Matter | 254,730.55 | 262,041.61 ± 19,977.02 | 251,688.00 | 257,489.23 ± 19,490.09 |
| Cerebral Cortex | 287,822.56 | 312,254.77 ± 21,819.79 | 285,806.03 | 314,283.32 ± 21,799.79 |
| Lateral Ventricle | 4,863.49 | 5,275.35 ± 2,989.19 | 5,133.03 | 5,472.29 ± 2,911.49 |
| Inferior Lateral Ventricle | 397.50 | 329.54 ± 70.31 | 440.80 | 393.61 ± 96.58 |
| Cerebellum White Matter | 15,975.83 | 16,301.85 ± 1,333.58 | 15,262.71 | 15,496.48 ± 1,279.35 |
| Cerebellum Cortex | 61,247.41 | 63,353.12 ± 4,984.67 | 61,222.93 | 63,149.82 ± 4,805.63 |
| Thalamus | 8,155.40 | 8,122.9 ± 620.73 | 8,588.93 | 8,677.38 ± 741.26 |
| Caudate | 4,581.43 | 4,463.68 ± 413.42 | 4,777.76 | 4,689.35 ± 407.37 |
| Putamen | 6,378.70 | 6,359.9 ± 478.71 | 6,281.82 | 6,291.88 ± 496.55 |
| Pallidum | 1,816.53 | 1,789.71 ± 151.18 | 2,057.41 | 2,028.37 ± 165.41 |
| Hippocampus | 4,503.63 | 4,472.23 ± 321.42 | 4,439.71 | 4,392.87 ± 319.81 |
| Amygdala | 2,088.20 | 2,076.29 ± 162.52 | 2,392.58 | 2,278.2 ± 181.43 |

## ACKNOWLEDGEMENT

We sincerely thank our participants and their whānau (families) for their time, trust, and contribution to this research. We also gratefully acknowledge the support of Gisborne Boys’ High School and Tūranga Health throughout this project. We are grateful to Mātai Ngā Māngai Māori for their guidance, to researchers and staff from Mātai Medical Research Institute and to our research participants for dedicating their time towards this study. We acknowledge the support extended by the Mātai mTBI Research Group, composed of Leigh Potter, Paul Condron, Davidson Taylor, Daniel Cornfield, Patrick McHugh, Taylor Emsden, Helen Danesh-Meyer, Matthew MacDonald, Gil Newburn, Tuterangi Nepe-Apatu, and Graeme Bydder. We would like to acknowledge the support of GE Healthcare for assistance with the MRI protocol.

## AUTHOR CONTRIBUTION STATEMENT

**Christian John Saludar:** Conceptualisation, Data curation, Investigation, Formal analysis, Writing-original draft, Writing-review and editing, Validation. **Maryam Tayebi:** Conceptualisation, Data curation, Investigation, Supervision, Validation, Writing-review and editing. **Eryn Kwon:** Data curation, Writing – review and editing, Funding acquisition, Project administration, Supervision. **Joshua McGeown:** Data curation, Writing – review and editing, Funding acquisition, Project administration. **Jacob Mathew:** Investigation, Writing-review and editing, Investigation. **William Schierding:** Validation, Writing-review and editing. **Mātai mTBI Research Group:** Data curation, Project administration, Resources. **Alan Wang:** Writing – review and editing, Supervision. **Justin Fernandez:** Writing – review and editing, Supervision. **Samantha Holdsworth:** Writing – review and editing, Supervision, Funding acquisition, Project administration. **Vickie Shim:** Writing – review and editing, Conceptualisation, Investigation, Supervision, Funding acquisition, Validation

## GLOSSARY

CSF: Cerebrospinal Fluid
dMRI: Diffusion MRI
FE: Finite Element
FOD: Fibre Orientation Distribution
GM: Gray matter
GOH: Gasser, Ogden, and Holzapfel
HAE: Head Acceleration Exposure
iMG: Instrumented Mouthguard
MRI: Magnetic Resonance Imaging
mTBI: mild Traumatic Brain Injury
TBI: Traumatic Brain Injury
WM: White matter

## TRANSPARENCY, RIGOR, AND REPRODUCIBILITY SUMMARY

This study is part of an ongoing longitudinal study in New Zealand. All procedures conducted in this study are in accordance with the ethics approval from the New Zealand Health and Disability Ethics Committee (20/NTB/14). All participants aged 16 and older provided informed consent, while participants under 16 provided assent with parental consent. For this study, general exclusion criteria included contraindication to MRI, neurological/psychiatric conditions, and dental braces affecting imaging quality.

A total of 78 male high school rugby players (aged 14-18 years old) participated in this study. Inclusion criteria required no mTBI within the past six months prior to start of study, no history of mTBI incident with loss of consciousness, no neurological disorders, no history of drug or excessive alcohol use and no diagnosis of dementia or delirium. Each scan included a multi-parametric MRI scan (*i.e.* structural, diffusion, functional MRI), and a cognitive and symptom assessment. More details of the parameters and tests used are reported in the manuscript. To record head acceleration exposure across the whole season, an instrumented mouthguard was provided for each rugby player.

A control group (14-18 years old) composed of non-collision sport, male athletes, was recruited and scanned at a single timepoint following the same protocol as the rugby players. The same inclusion and exclusion criteria were applied for the control group, with the addition of no self-reported mTBI history or participation in collision sports within the past two years.

The primary aim of this study is to establish a computational framework that enables the creation of an average brain from MRI scans of subjects and to develop an FE model. Moreso, this FE model will incorporate fibre dispersion parameter from diffusion MRI and be validated against human head cadaveric experiments reported in the literature. This study is among the few to present a complete framework from MRI to finite element modelling using open-access tools, making it reproducible.

## DATA AVAILABILITY

In accordance with our Indigenous and community engagement policies, non-identifiable data are available upon request and subsequent approval by the Mātai Ngā Māngai Māori Board (contacted at)

## REFERENCES

1. Zhao W, Ji S. Displacement-and Strain-Based Discrimination of Head Injury Models across a Wide Range of Blunt Conditions. Ann Biomed Eng. 2020;48:1661–77. 10.1007/s10439-020-02496-y

2. Sbriglio C, Ptak M, Kwiatkowski A. Advances in Computational Modelling of Head Injury Biomechanics – a Comprehensive Review. Archives of Computational Methods in Engineering. 2025; 10.1007/s11831-025-10463-w

3. Ji S, Zhao W, Ford JC, Beckwith JG, Bolander RP, Greenwald RM, et al. Group-Wise Evaluation and Comparison of White Matter Fiber Strain and Maximum Principal Strain in Sports-Related Concussion. J Neurotrauma. 2015;32:441–54. 10.1089/neu.2013.3268

4. Mao H, Zhang L, Jiang B, Genthikatti V V., Jin X, Zhu F, et al. Development of a Finite Element Human Head Model Partially Validated With Thirty Five Experimental Cases. J Biomech Eng. 2013;135. 10.1115/1.4025101

5. Miller LE, Urban JE, Stitzel JD. Development and validation of an atlas-based finite element brain model. Biomech Model Mechanobiol. 2016;15:1201–14. 10.1007/s10237-015-0754-1

6. Kleiven S, von Holst H. Consequences of head size following trauma to the human head. J Biomech. 2002;35:153–60. 10.1016/S0021-9290(01)00202-0

7. Li X. Subject-Specific Head Model Generation by Mesh Morphing: A Personalization Framework and Its Applications. Front Bioeng Biotechnol. 2021;9. 10.3389/fbioe.2021.706566

8. Escarcega JD, Okamoto RJ, Alshareef AA, Johnson CL, Bayly P V. Effects of anatomy and head motion on spatial patterns of deformation in the human brain. Ann Biomed Eng. 2025;53:867–80. 10.1007/s10439-024-03671-1

9. Zou Y, Zhu W, Yang H-C, Jang I, Vike NL, Svaldi DO, et al. Development of brain atlases for early-to-middle adolescent collision-sport athletes. Sci Rep. 2021;11:6440. 10.1038/s41598-021-85518-6

10. Shim V, Tayebi M, Kwon E, Guild S-J, Scadeng M, Dubowitz D, et al. Combining advanced magnetic resonance imaging (MRI) with finite element (FE) analysis for characterising subject-specific injury patterns in the brain after traumatic brain injury. Eng Comput. 2022;38:3925–37. 10.1007/s00366-022-01697-4

11. Dewan MC, Rattani A, Gupta S, Baticulon RE, Hung Y-C, Punchak M, et al. Estimating the global incidence of traumatic brain injury. J Neurosurg. 2019;130:1080–97. 10.3171/2017.10.JNS17352

12. Te Ao B, Brown P, Tobias M, Ameratunga S, Barker-Collo S, Theadom A, et al. Cost of traumatic brain injury in New Zealand. Neurology. 2014;83:1645–52. 10.1212/WNL.0000000000000933

13. Feigin VL, Theadom A, Barker-Collo S, Starkey NJ, McPherson K, Kahan M, et al. Incidence of traumatic brain injury in New Zealand: a population-based study. Lancet Neurol. 2013;12:53–64. 10.1016/S1474-4422(12)70262-4

14. Silverberg ND, Iverson GL, Cogan A, Dams-O-Connor K, Delmonico R, Graf MJP, et al. The American Congress of Rehabilitation Medicine Diagnostic Criteria for Mild Traumatic Brain Injury. Arch Phys Med Rehabil. 2023;104:1343–55. 10.1016/j.apmr.2023.03.036

15. Davenport EM, Whitlow CT, Urban JE, Espeland MA, Jung Y, Rosenbaum DA, et al. Abnormal white matter integrity related to head impact exposure in a season of high school varsity football. J Neurotrauma. 2014;31:1617–24. 10.1089/neu.2013.3233

16. Tayebi M, Kwon E, McGeown J, Potter L, Taylor D, Condron P, et al. Characterizing the Effect of Repetitive Head Impact Exposure and mTBI on Adolescent Collision Sports Players’ Brain with Diffusion Magnetic Resonance Imaging. J Neurotrauma. 2024; 10.1089/neu.2024.0064

17. Hume PA, Theadom A, Lewis GN, Quarrie KL, Brown SR, Hill R, et al. A Comparison of Cognitive Function in Former Rugby Union Players Compared with Former Non-Contact-Sport Players and the Impact of Concussion History. Sports Medicine. 2017;47:1209–20. 10.1007/s40279-016-0608-8

18. Bellomo G, Piscopo P, Corbo M, Pupillo E, Stipa G, Beghi E, et al. A systematic review on the risk of neurodegenerative diseases and neurocognitive disorders in professional and varsity athletes. Neurol Sci. 2022;43:6667–91. 10.1007/s10072-022-06319-x

19. McGeown JP, Pedersen M, Mito R, Theadom A, Maller JJ, Condron P, et al. Neuroimaging correlates of symptom burden and functional recovery following mild traumatic brain injury: A systematic review. Neuroimage Clin. 2026;49:103910. 10.1016/j.nicl.2025.103910

20. Carmo GP, Dymek M, Ptak M, Alves-de-Sousa RJ, Fernandes FAO. Development, validation and a case study: The female finite element head model (FeFEHM). Comput Methods Programs Biomed. 2023;231:107430. 10.1016/j.cmpb.2023.107430

21. Shim VB, Holdsworth S, Champagne AA, Coverdale NS, Cook DJ, Lee T-R, et al. Rapid Prediction of Brain Injury Pattern in mTBI by Combining FE Analysis With a Machine-Learning Based Approach. IEEE Access. 2020;8:179457–65. 10.1109/ACCESS.2020.3026350

22. Ghajari M, Hellyer PJ, Sharp DJ. Computational modelling of traumatic brain injury predicts the location of chronic traumatic encephalopathy pathology. Brain. 2017;140:333–43. 10.1093/brain/aww317

23. McGeown JP, Pedersen M, Bussey M, Schierding WS, Condron P, Emsden T, et al. Longitudinal changes in network-based functional connectivity over a rugby season in adolescent males. 2025. 10.1101/2025.03.16.25324069

24. Holm L, Cassidy JD, Carroll LJ, Borg J, Neurotrauma Task Force on Mild Traumatic Brain Injury of the WHO Collaborating Centre. Summary of the WHO Collaborating Centre for Neurotrauma Task Force on Mild Traumatic Brain Injury. J Rehabil Med. 2005;37:137–41. 10.1080/16501970510027321

25. Stitt D, Draper N, Alexander K, Kabaliuk N. Laboratory Validation of Instrumented Mouthguard for Use in Sport. Sensors. 2021;21:6028. 10.3390/s21186028

26. Field B, Waddington G, McKune A, Goecke R, Gardner AJ. Validation of an instrumented mouthguard in rugby union—a pilot study comparing impact sensor technology to video analysis. Front Sports Act Living. 2023;5. 10.3389/fspor.2023.1230202

27. Isensee F, Schell M, Pflueger I, Brugnara G, Bonekamp D, Neuberger U, et al. Automated brain extraction of multisequence MRI using artificial neural networks. Hum Brain Mapp. 2019;40:4952–64. 10.1002/hbm.24750

28. Gaser C, Dahnke R, Thompson PM, Kurth F, Luders E, the Alzheimer’s Disease Neuroimaging Initiative. CAT: a computational anatomy toolbox for the analysis of structural MRI data. Gigascience. 2024;13. 10.1093/gigascience/giae049

29. Tustison NJ, Avants BB, Cook PA, Yuanjie Zheng, Egan A, Yushkevich PA, et al. N4ITK: Improved N3 Bias Correction. IEEE Trans Med Imaging. 2010;29:1310–20. 10.1109/TMI.2010.2046908

30. Woolrich MW, Jbabdi S, Patenaude B, Chappell M, Makni S, Behrens T, et al. Bayesian analysis of neuroimaging data in FSL. Neuroimage. 2009;45:S173–86. 10.1016/j.neuroimage.2008.10.055

31. Smith SM, Jenkinson M, Woolrich MW, Beckmann CF, Behrens TEJ, Johansen-Berg H, et al. Advances in functional and structural MR image analysis and implementation as FSL. Neuroimage. 2004;23:S208–19. 10.1016/j.neuroimage.2004.07.051

32. Tournier J-D, Smith R, Raffelt D, Tabbara R, Dhollander T, Pietsch M, et al. MRtrix3: A fast, flexible and open software framework for medical image processing and visualisation. Neuroimage. 2019;202:116137. 10.1016/j.neuroimage.2019.116137

33. Veraart J, Novikov DS, Christiaens D, Ades-aron B, Sijbers J, Fieremans E. Denoising of diffusion MRI using random matrix theory. Neuroimage. 2016;142:394–406. 10.1016/j.neuroimage.2016.08.016

34. Andersson JLR, Sotiropoulos SN. An integrated approach to correction for off-resonance effects and subject movement in diffusion MR imaging. Neuroimage. 2016;125:1063–78. 10.1016/j.neuroimage.2015.10.019

35. Schilling KG, Blaber J, Huo Y, Newton A, Hansen C, Nath V, et al. Synthesized b0 for diffusion distortion correction (Synb0-DisCo). Magn Reson Imaging. 2019;64:62–70. 10.1016/j.mri.2019.05.008

36. Bastiani M, Cottaar M, Fitzgibbon SP, Suri S, Alfaro-Almagro F, Sotiropoulos SN, et al. Automated quality control for within and between studies diffusion MRI data using a non-parametric framework for movement and distortion correction. Neuroimage. 2019;184:801–12. 10.1016/j.neuroimage.2018.09.073

37. Jenkinson M, Beckmann CF, Behrens TEJ, Woolrich MW, Smith SM. FSL. Neuroimage. 2012;62:782–90. 10.1016/j.neuroimage.2011.09.015

38. Avants BB, Tustison NJ, Song G, Cook PA, Klein A, Gee JC. A reproducible evaluation of ANTs similarity metric performance in brain image registration. Neuroimage. 2011;54:2033–44. 10.1016/j.neuroimage.2010.09.025

39. Giordano C, Zappalà S, Kleiven S. Anisotropic finite element models for brain injury prediction: the sensitivity of axonal strain to white matter tract inter-subject variability. Biomech Model Mechanobiol. 2017;16:1269–93. 10.1007/s10237-017-0887-5

40. Dhollander T, Mito R, Raffelt D, Connelly A. Improved white matter response function estimation for 3-tissue constrained spherical deconvolution. 2019.

41. Billot B, Greve DN, Puonti O, Thielscher A, Van Leemput K, Fischl B, et al. SynthSeg: Segmentation of brain MRI scans of any contrast and resolution without retraining. Med Image Anal. 2023;86:102789. 10.1016/j.media.2023.102789

42. Desikan RS, Ségonne F, Fischl B, Quinn BT, Dickerson BC, Blacker D, et al. An automated labeling system for subdividing the human cerebral cortex on MRI scans into gyral based regions of interest. Neuroimage. 2006;31:968–80. 10.1016/j.neuroimage.2006.01.021

43. Wasserthal J, Neher P, Maier-Hein KH. TractSeg - Fast and accurate white matter tract segmentation. Neuroimage. 2018;183:239–53. 10.1016/j.neuroimage.2018.07.070

44. Winkler AM, Ridgway GR, Webster MA, Smith SM, Nichols TE. Permutation inference for the general linear model. Neuroimage. 2014;92:381–97. 10.1016/j.neuroimage.2014.01.060

45. Fedorov A, Beichel R, Kalpathy-Cramer J, Finet J, Fillion-Robin J-C, Pujol S, et al. 3D Slicer as an image computing platform for the Quantitative Imaging Network. Magn Reson Imaging. 2012;30:1323–41. 10.1016/j.mri.2012.05.001

46. Maas SA, Ellis BJ, Ateshian GA, Weiss JA. FEBio: Finite Elements for Biomechanics. J Biomech Eng. 2012;134. 10.1115/1.4005694

47. Shim VB, Fernandez JW, Gamage PB, Regnery C, Smith DW, Gardiner BS, et al. Subject-specific finite element analysis to characterize the influence of geometry and material properties in Achilles tendon rupture. J Biomech. 2014;47:3598–604. 10.1016/j.jbiomech.2014.10.001

48. Giordano C, Kleiven S. Connecting fractional anisotropy from medical images with mechanical anisotropy of a hyperviscoelastic fibre-reinforced constitutive model for brain tissue. J R Soc Interface. 2014;11. 10.1098/rsif.2013.0914

49. Kleiven S. Evaluation of head injury criteria using a finite element model validated against experiments on localized brain motion, intracerebral acceleration, and intracranial pressure. International Journal of Crashworthiness. 2006;11:65–79. 10.1533/ijcr.2005.0384

50. Giordano C, Cloots RJH, van Dommelen JAW, Kleiven S. The influence of anisotropy on brain injury prediction. J Biomech. 2014;47:1052–9. 10.1016/j.jbiomech.2013.12.036

51. Miller RT, Margulies SS, Leoni M, Nonaka M, Chen X, Smith DH, et al. Finite Element Modeling Approaches for Predicting Injury in an Experimental Model of Severe Diffuse Axonal Injury. 1998. 10.4271/983154

52. Alshareef A, Giudice JS, Forman J, Salzar RS, Panzer MB. A Novel Method for Quantifying Human *In Situ* Whole Brain Deformation under Rotational Loading Using Sonomicrometry. J Neurotrauma. 2018;35:780–9. 10.1089/neu.2017.5362

53. Hardy WN, Mason MJ, Foster CD, Shah CS, Kopacz JM, Yang KH, et al. A Study of the Response of the Human Cadaver Head to Impact. 2007. 10.4271/2007-22-0002

54. Gehre C, Gades H, Wernicke P. Objective rating of signals using test and simulation responses. 2009. https://api.semanticscholar.org/CorpusID:107307960

55. Fernandez JW, Mithraratne P, Thrupp SF, Tawhai MH, Hunter PJ. Anatomically based geometric modelling of the musculo-skeletal system and other organs. Biomech Model Mechanobiol. 2004;2:139–55. 10.1007/s10237-003-0036-1

56. Haynes W. Benjamini–Hochberg Method. Encyclopedia of Systems Biology. New York, NY: Springer New York; 2013. p. 78–78. 10.1007/978-1-4419-9863-7_1215

57. Cohen J. Statistical Power Analysis. Curr Dir Psychol Sci. 1992;1:98–101. 10.1111/1467-8721.ep10768783

58. Pfister T, Pfister K, Hagel B, Ghali WA, Ronksley PE. The incidence of concussion in youth sports: a systematic review and meta-analysis. Br J Sports Med [Internet]. 2016;50:292. 10.1136/bjsports-2015-094978

59. Tayebi M, Holdsworth SJ, Champagne AA, Cook DJ, Nielsen P, Lee T-R, et al. The role of diffusion tensor imaging in characterizing injury patterns on athletes with concussion and subconcussive injury: a systematic review. Brain Inj. 2021;35:621–44. 10.1080/02699052.2021.1895313

60. Koerte IK, Wiegand TLT, Bonke EM, Kochsiek J, Shenton ME. Diffusion Imaging of Sport-related Repetitive Head Impacts—A Systematic Review. Neuropsychol Rev. 2023;33:122–43. 10.1007/s11065-022-09566-z

61. Lees B, Earls NE, Meares S, Batchelor J, Oxenham V, Rae CD, et al. Diffusion Tensor Imaging in Sport-Related Concussion: A Systematic Review Using an *a priori* Quality Rating System. J Neurotrauma. 2021;38:3032–46. 10.1089/neu.2021.0154

62. Wilke M, Schmithorst VJ, Holland SK. Assessment of spatial normalization of whole-brain magnetic resonance images in children. Hum Brain Mapp. 2002;17:48–60. 10.1002/hbm.10053

63. Mazziotta JC, Toga AW, Evans A, Fox P, Lancaster J. A Probabilistic Atlas of the Human Brain: Theory and Rationale for Its Development. Neuroimage. 1995;2:89–101. 10.1006/nimg.1995.1012

64. Atilano-Barbosa D, Barrios FA. Brain morphological variability between whites and African Americans: the importance of racial identity in brain imaging research. Front Integr Neurosci. 2023;17:1027382. 10.3389/fnint.2023.1027382

65. Ota K, Nakazato Y, Oyama G. Developing and validating an elderly brain template: A comprehensive comparison with MNI152 for age-specific neuroimaging analyses. Neuroimage. 2025;320:121473. 10.1016/j.neuroimage.2025.121473

66. Miller LE, Urban JE, Espeland MA, Walkup MP, Holcomb JM, Davenport EM, et al. Cumulative strain-based metrics for predicting subconcussive head impact exposure-related imaging changes in a cohort of American youth football players. J Neurosurg Pediatr. American Association of Neurological Surgeons; 2022;29:387–96. 10.3171/2021.10.PEDS21355

67. Giudice JS, Zeng W, Wu T, Alshareef A, Shedd DF, Panzer MB. An Analytical Review of the Numerical Methods used for Finite Element Modeling of Traumatic Brain Injury. Ann Biomed Eng. 2019;47:1855–72. 10.1007/s10439-018-02161-5

68. Ji S, Ghajari M, Mao H, Kraft RH, Hajiaghamemar M, Panzer MB, et al. Use of Brain Biomechanical Models for Monitoring Impact Exposure in Contact Sports. Ann Biomed Eng. 2022;50:1389–408. 10.1007/s10439-022-02999-w

69. Jenkinson M. Non-linear registration aka Spatial normalisation. 2007. https://api.semanticscholar.org/CorpusID:16693956

70. Maas SA, Ellis BJ, Ateshian GA, Weiss JA. FEBio: Finite Elements for Biomechanics. J Biomech Eng. 2012;134. 10.1115/1.4005694

71. Li X, Zhou Z, Kleiven S. An anatomically detailed and personalizable head injury model: Significance of brain and white matter tract morphological variability on strain. Biomech Model Mechanobiol. 2021;20:403–31. 10.1007/s10237-020-01391-8

72. Zhao W, Choate B, Ji S. Material properties of the brain in injury-relevant conditions – Experiments and computational modeling. J Mech Behav Biomed Mater. 2018;80:222–34. 10.1016/j.jmbbm.2018.02.005

73. Shim VB, Battley M, Anderson IA, Munro JT. Validation of an efficient method of assigning material properties in finite element analysis of pelvic bone. Comput Methods Biomech Biomed Engin. 2015;18:1495–9. 10.1080/10255842.2014.920831

74. Hiscox L V., McGarry MDJ, Schwarb H, Van Houten EEW, Pohlig RT, Roberts N, et al. Standard-space atlas of the viscoelastic properties of the human brain. Hum Brain Mapp. 2020;41:5282–300. 10.1002/hbm.25192

75. Knutsen AK, Gomez AD, Gangolli M, Wang W-T, Chan D, Lu Y-C, et al. In vivo estimates of axonal stretch and 3D brain deformation during mild head impact. Brain Multiphys. 2020;1:100015. 10.1016/j.brain.2020.100015

76. Upadhyay K, Alshareef A, Knutsen AK, Johnson CL, Carass A, Bayly P V., et al. Development and validation of subject-specific 3D human head models based on a nonlinear visco-hyperelastic constitutive framework. J R Soc Interface. 2022;19. 10.1098/rsif.2022.0561

77. Alshareef A, Carass A, Lu Y-C, Mojumder J, Diano AM, Bailey OM, et al. Average Biomechanical Responses of the Human Brain Grouped by Age and Sex. Ann Biomed Eng. 2025;53:1496–511. 10.1007/s10439-025-03725-y

